# Computational Phenotyping of Treatment-Resistant Depression prior to Electroconvulsive Therapy

**DOI:** 10.1101/2024.10.02.24314360

**Authors:** Rachel E. Jones, L. Paul Sands, Jonathan D. Trattner, Angela Jiang, Christina K. Johnson, Evelyn B. Farkas, Predrag V. Gligorovic, Heather E. Douglas, Rommel Ramos, Kenneth T. Kishida

## Abstract

**KEY POINTS:** **QUESTION**: Can neurocomputational depictions of learning and affective behavior characterize patients with treatment-resistant depression before electroconvulsive therapy?

**FINDINGS**: In this observational study, computational models were used to quantify the behavioral dynamics of 1) adaptive choice behavior as individuals learned from feedback and 2) associated changes in affective self-report. These models provided quantitative parameters that were associated with specific neural and behavioral changes in patients with treatment-resistant depression and may be sufficient to independently identify patients with depression.

**MEANING**: Computational models that describe hypothesized mechanisms underlying adaptive behavior and affective experience may provide a means to quantitatively phenotype individual differences in major depression pathophysiology.

**IMPORTANCE:** Globally, treatment-resistant depression affects approximately one-third of all patients diagnosed with major depressive disorder. Currently, there are neither behavioral nor neural measures that quantitatively phenotype characteristics underlying treatment-resistant depression.

**OBJECTIVE:** Determine whether neurocomputational models that integrate information about adaptive behavior and associated self-reported feelings can characterize differences in patients with treatment-resistant depression.

**DESIGN:** In this observational study, data were collected over two research visits from 2020-2023 that occurred before and after standard-of-care electroconvulsive therapy (ECT) for treatment-resistant depression. This report focuses on “visit 1”, which occurred after patients consented to ECT but before their initial treatment.

**SETTING:** Wake Forest University School of Medicine; Atrium Health Wake Forest Baptist Psychiatric Outpatient Center; Atrium Health Wake Forest Hospital.

**PARTICIPANTS:** Participants planning to receive ECT for depression (“pre-ECT”) and participants not planning to receive ECT with (“non-ECT”) or without depression (“no-depression”), were recruited from the Psychiatric Outpatient Center and community.

**EXPOSURES:** Computerized delivery of a ‘Probabilistic Reward and Punishment with Subjective Rating’ task during fMRI.

**MAIN OUTCOMES AND MEASURES:** Computational modeling of choice behavior provided parameters that characterized learning dynamics and associated affect dynamics expressed through intermittent Likert scale self-reports. Multivariate statistical analyses relating model parameters, neurobehavioral responses, and clinical assessments.

**RESULTS:** Pre-ECT (N=29; 55.2% female), non-ECT (N=40; 70% female), and no-depression (N=41; 65.9% female). Parameters derived from computational models fit to behavior elicited during learning *and* the expression of affective experiences clearly differentiates the three groups. Reinforcement Learning model parameters alone do not perform as well as models that incorporate affective self-reports. Notably, the set of model parameters that include learning *and* affective dynamics demonstrated excellent, cross-validated, diagnostic classification of depression diagnosis. Prior to ECT, neurobehavioral responses associated with learning and affective experiences about ‘punishing’ events were significantly impaired in pre-ECT compared to non-ECT and no-depression cohorts.

**CONCLUSIONS AND RELEVANCE:** Computational models of behavioral dynamics associated with learning and affect can describe specific hypotheses about neurocomputational-mechanisms underlying treatment-resistant depression. The present work suggests differences in processing of emotionally negative states and suggests a potential model-based behavioral diagnostic for individuals with major depression. Such models may eventually be used to augment the diagnosis of treatment-resistant depression or possibly determine phenotype-genotype relationships for disease status and progression.

## INTRODUCTION

Depression is projected to be the leading cause of global disability by 2030^1^ with approximately one-third of patients meeting criteria for treatment-resistant depression (TRD)^2,3^. These patients suffer for prolonged periods before effective interventions such as electroconvulsive therapy (ECT) may be performed^4,5^. Unbiased neurobehavioral measures that identify patients at risk for TRD may be leveraged to accelerate research into the neurobehavioral mechanisms underlying depression, which in turn may lead to earlier diagnosis and treatment strategies^6,7^.

Recent work suggests *computational psychiatric* methods may aid in investigating depression pathophysiology^8–10^. Notably, computational reinforcement learning (RL) models have been used to reveal connections between moment-to-moment decisions, neural responses, momentary ‘happiness’, and depression symptoms^11–14^. However, little computational psychiatric work has been done in TRD. Here, we investigated mechanisms underlying TRD using functional magnetic resonance imaging (fMRI), a value-based decision-making task, and RL-based computational models describing 1) hypothesized reward and punishment learning mechanisms and 2) associated affective behavior. This study uses a Valence Partitioned Reinforcement Learning framework^15^ that was recently shown to provide a good explanation of behavior^16,17^ and track sub-second changes in dopamine levels^16^ in the task used in the present work.

Symptoms of depression have been linked to changes in processing of both positive and negative valence^18–21^. Changes in positive valence processing may be associated with anhedonia, while changes in negative valence processing may relate to feelings of hopelessness or worthlessness, difficulties in regulating negative emotions, or negative rumination^22–24^. Research consistently shows that *reward prediction error* (RPE)^25,26^-associated brain responses are reduced in patients with depression^27–29^. Such findings broadly suggest that observable learning behavior and computational models that connect affective responses to reward processing may be fruitfully used to investigate depression^13,30–34^. Collectively, computational models of learning and affect have consistently identified disruptions in reward processing in depression; but, work investigating the impact of negative valence is less coherent^35–38^.

RL model based studies report contradictory changes in neural activity associated with punishment learning in patients with depression^39–42^. Apparent inconsistencies may stem from the use of RL models that depict punishments as symmetric reflections of rewards rather than as potentially separate, independently processed signals^15,43–45^. Valance-partitioned approaches have been suggested in the RL literature whereby the expectations of rewards and punishments may be learned at different rates^15–17,46–48^. Modeling learning and affective behavior through such a framework may clarify important distinctions about how negative and positive experiences differentially impact patients with depression. For example, Brown and colleagues^12^ demonstrated that separating RL model parameters based on the valence of each trial better explained both control and depression participant choice behavior compared to methods that did not discriminate trial valence^12^.

Here, we follow this line of reasoning and, using a previously validated Valence Partitioned Reinforcement Learning (VPRL) model^15–17^, sought to determine whether a computational psychiatric approach^50–52^ that partitions positive and negative valence and utilizes derived learning signals to predict affective responses could provide insight into patients with TRD. We hypothesized that individual differences in depression may be revealed in both VPRL parameters and model parameters that describe how latent learning signals modulate affective self-reports.

## METHODS

### Study Design

Here, we report data collected during research “visit 1” of a two-visit observational study following a standard-of-care ECT-treatment timeline (Fig. 1A). All participants completed a Probabilistic Reward and Punishment with Subjective Rating task^16,17^ with fMRI scanning. The Patient Health Questionnaire 9 (PHQ-9^49^), Hamilton Depression Rating Scale (HAM-D^50^), and Montreal Cognitive Assessment (MOCA^51^) were assessed after fMRI scanning. Participants provided written informed consent under IRB protocol: WFUSoM:IRB00056131.

**Fig. 1.**
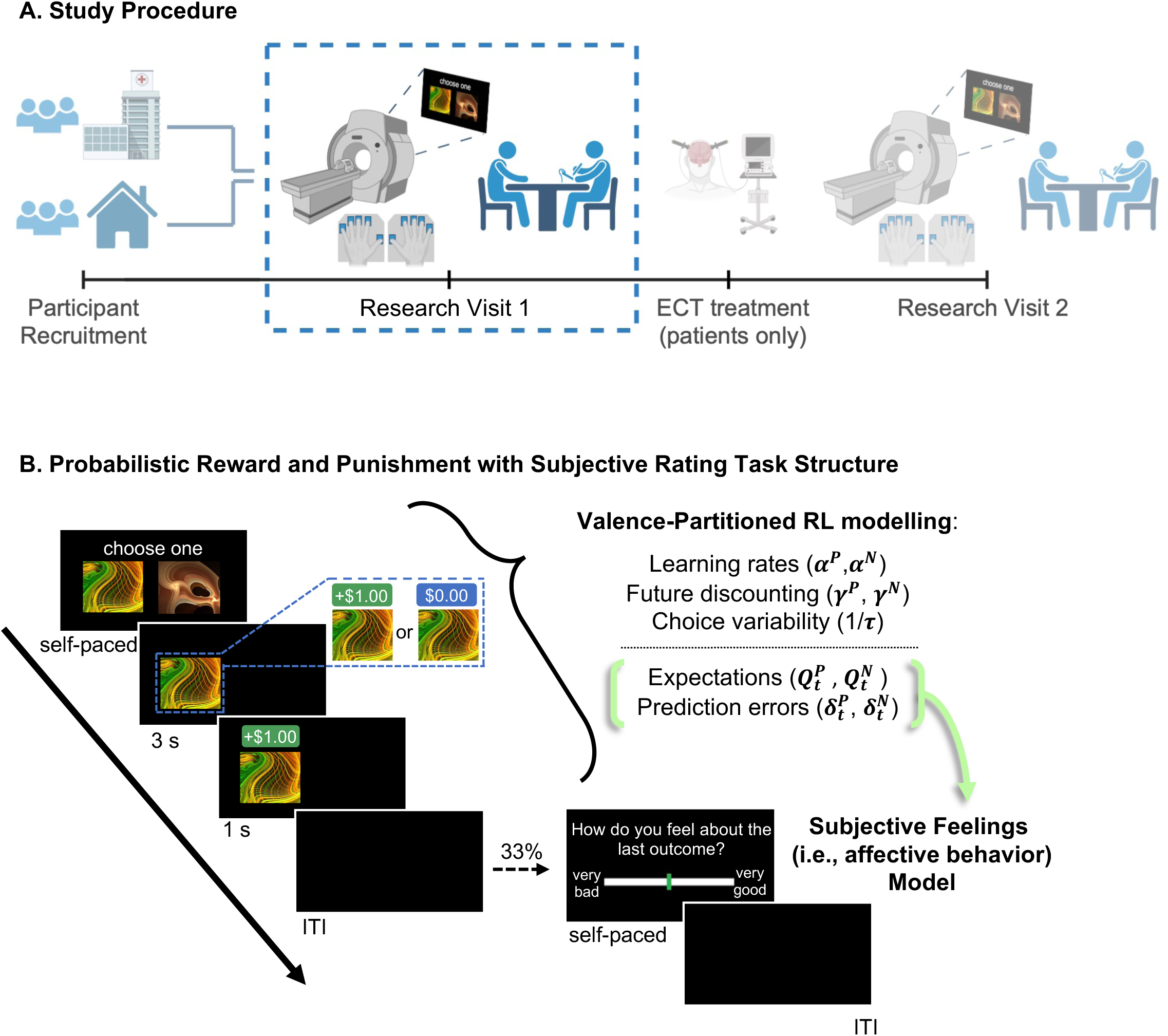
Study procedure and PRPwSR task structure. **(A)** Patients were recruited from AHWFB Psychiatry & Behavioral Health outpatient clinic. Community volunteers with and without depression were also recruited. Participants complete a probabilistic reward and punishment with subjective rating (PRPwSR) task with fMRI scanning, followed by clinical assessment for each research visit. Here, we focus on research visit 1 (pre-ECT treatment). **(B)** Schematic of a trial in the PRPwSR task. Each trial starts with an option presentation screen. A participant selects an option (self-paced), and the other option disappears. After 3s, the chosen option is reinforced probabilistically (e.g., the selected win icon is associated with a certain probability of winning $1 versus $0), and the outcome is shown for 1s. The screen then presents an interstimulus interval (ITI - sampled from a Poison distribution with =3 seconds). After each trial, there is a 33% probability of a rating screen that asks participants how they felt about their most recent outcome. We fit a valence-partitioned reinforcement learning model to participants’ choice behavior in the PRPwSR task. This produced learning parameter estimates (*α^P^, α^N^, γ^P^, γ^N^*, and *τ*) that we used to calculate participants’ expectations 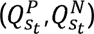 and prediction errors 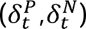 during the subjective rating trials. Participants’ expectations and prediction errors were then used as independent predictors of their reported ratings in a Subjective Feeling linear regression model to relate learning signals to affective experience (i.e., green brackets and arrow).

### Participants

“Pre-ECT” patients included patients with TRD who consented to begin, but were naïve to, ECT at AHWFB Psychiatric Outpatient Center (pre-ECT,N=29; 55.17% female). Participants with depression not planning ECT (non-ECT,N=40; 70% female) and participants without depression (no-depression,N=41; 65.90% female) were recruited from the Winston-Salem, NC area. See eMethods and eTables1-2 for full details.

### Probabilistic Reward and Punishment with Subjective Rating (PRPwSR) Task

The PRPwSR task (Fig. 1B, eMethods) was executed as previously described^16,17^. Briefly, the PRPwSR task is a value-based decision-making task. Over 150 trials, participants make choices, experience monetary gains and losses, and must learn which probabilistic options maximize real monetary returns (eFigs.1-3). After each trial, with one-third probability, participants are asked, “How do you feel about the last outcome?”. Participants respond using a Likert scale ranging “very bad” to “very good.”

### Computational Modeling

We used hierarchical Bayesian methods^52^ to fit models to participants’ behavior on the PRPwSR task (eMethods). Behavior was modeled as a state-action-outcome learning problem consistent with computational RL theory. Expected values and outcome prediction errors were estimated using a VPRL framework^15–17^.

These signals were used to model and predict subjective ratings^17^. Each individual was represented as a vector of their unique set of parameters for further analyses, which we hypothesized may represent a *computational phenotype*^53,54^. A description of parameters follows:

“Valence Partitioned Reinforcement Learning” parameters

A previously validated VPRL framework^15–17^ was used to summarize participants’ behavior. The mathematical description of this VPRL model^15–17^ is described (eMethods). Here, we describe parameters in narrative form for interpretability:

VPRL^15^ expresses the hypothesis that brains track appetitive (e.g., positive/rewarding) and aversive (e.g., negative/punishing) stimuli simultaneously, but via independent systems. This allows stimuli to predict benefits and/or costs that may be independently estimated such that cost-benefit comparisons can be made^15^. Each system updates *value estimates* (i.e., Q-values) akin to standard Q-learning with temporal difference RL rules^45,55^. Hence, Q-values for both positive 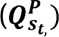 and negative 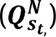 states (s_t_) are estimated. Likewise, future states’ (*s_t+1_*) positive 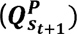 and negative 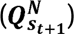 values are estimated and, respectively, discounted by parameters (*γ^P^*, *γ^N^*). The combination of current actual outcomes, discounted expectations of future values, and expected current values are used to calculate temporal difference prediction errors 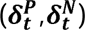, which are then used to update respective Q-values by independent learning weights (*α^P^*,*α^N^*). We modeled choice policy using a softmax function with a temperature parameter (τ) that describes how exploitative versus random a participant’s choices are given Q-value estimates. The free parameters in the VPRL framework (α^P^, α^N^, γ^P^, γ^N^, and τ) were used to characterize each participant’s computational *learning* phenotype.

“Subjective Feeling” Model parameters

We hypothesized that participants’ subjective ratings resulted from a combination of expectations 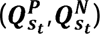 and better or worse than expected outcomes 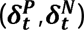. Further, we hypothesized 1) that expectations about the unchosen option (i.e., the counterfactual choice) may play a significant role in participants’ affective response; and 2) that the positive or negative sign of outcome prediction error – indicating greater-than or less-than expected, respectively – would be asymmetrically weighted in their influence on affective responses. Therefore, we fit a linear regression model with subjective ratings as the dependent variable and Q-values 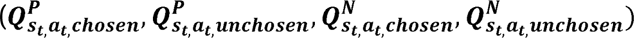 and signed prediction errors 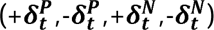 as independent variables. See eMethods for details. eFig.4 shows model performances.

### fMRI Analysis

eMethods contains details about fMRI data-acquisition, pre-processing, and model-based analyses. VPRL and Subjective Feeling models were fit to each participants’ behavior. Following first-level general linear modeling, results for each participant were included in a second-level whole-brain analysis of the variance (ANOVA) for group comparisons. Post-hoc t-tests were performed given significant ANOVA results. All comparisons were initially assessed at an uncorrected threshold of p<0.001 and reported results were selected using a family-wise error-corrected threshold of p<0.05 at cluster and peak voxel levels.

### Statistical Analyses

We used hierarchical Bayesian analysis^52^ to estimate posterior distributions of free parameters in the VPRL model of choice behavior and learning (*α^P^, α^N^, γ^P^, γ^N^*, and τ) and subsequently for the linear regression coefficients in the Subjective Feeling regression model 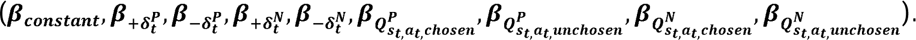. Differences across groups were assessed by considering differences between individual parameters and by performing a principal component analysis (PCA) to reduce the dimensionality of both parameter sets.

To test whether model-based summaries of behavior could be used to characterize individual differences without prior information about participants’ clinical diagnoses, we fit VPRL and Subjective Feeling models assuming no prior clinical information and uniform priors over parameter values. The resulting parameter estimates were concatenated to form a single 14-vector per participant and subjected to PCA. Finally, we performed a leave-one-out cross-validated linear discriminant analysis (LDA) to determine if the 14-vector could distinguish depression versus no depression from behavior observed in the PRPwSR task. Receiver Operating Characteristic (ROC) analyses are reported. Further details provided in eMethods. All analyses were conducted in R version 4.2.2 and RStan version 2.21.8^56^.

## RESULTS

### Clinical Characterization

Pre-ECT, non-ECT, and no-depression groups are significantly different on both PHQ-9 (F_2,107_=112) and HAM-D (F_2,107_=121.5). PHQ-9 and HAM-D scores are significantly higher for pre-ECT compared to non-ECT (post-hoc t-test, p<0.001) or no-depression (post-hoc t-test, p<0.001); non-ECT is significantly greater than no-depression (post-hoc t-tests, p<0.001). The three groups did not differ in age, gender, race, ethnicity, or MOCA score. eTables1-2 summarize participant demographic and clinical characteristics.

### Behavioral and Neural *Computational Phenotypes* for Reward and Punishment Learning – VPRL Behavioral

We hypothesized that VPRL parameters determined from participants’ behavior on the PRPwSR task would distinguish depression diagnosis and ECT treatment status. To test this hypothesis, we first determined the best groupings for model fitting using hierarchical Bayesian methods^57,58^. We compared VPRL model fits separately for (i) all participants as one group, (ii) depression (i.e., pre-ECT and non-ECT combined) and no-depression groupings; and (iii) pre-ECT, non-ECT, and no-depression separated. The latter, three-group model, resulted in the maximum model evidence and predictive density values (eTable3) and was used for further analyses (Fig. 2, Table 1).

**Table 1.** Group-level differences in BOLD responses to learning and subjective experience signals.

| Parameter | Region | Voxels | Cluster-level p-value | Peak-level p-value | Statistic | Peak MNI coordinates [x y z] |
| --- | --- | --- | --- | --- | --- | --- |
| **Learning Computations** |  |  |  |  |  |  |
| $+\delta_t^P$ | | | | | NS | |
| $-\delta_t^P$ | | | | | NS | |
| $Q_{s_t, a_t, chosen}^P$ | Right calcarine sulcus | 188 | **0.011**<br>**0.001**<br>**0.002** | 0.473<br>0.112<br>0.689 | F(2,101)=12.77<br><sup>a</sup>no-depression > pre-ECT<br><sup>a</sup>non-ECT > pre-ECT | [14 -74 16] |
| $Q_{s_t, a_t, unchosen}^P$ | | | | | NS | |
| $+\delta_t^N$ | Medial prefrontal cortex | 74 | **0.027**<br>**5.24e-6** | 0.998<br>0.992 | F(2,101)=9.37<br><sup>a</sup>non-ECT > no-depression | [4 54 16] |
|  | Ventromedial prefrontal cortex | 186 | **9.20e-5**<br>**1.78e-8** | 0.526<br>0.141 | F(2,101)=13.01<br><sup>a</sup>pre-ECT > no-depression | [0 58 -2] |
| $-\delta_t^N$ | Left supplementary motor area | 70 | **0.020**<br>**6.72e-5** | 0.096<br>**1.73e-4** | F(2,101)=15.70<br><sup>a</sup>pre-ECT > no-depression | [-4 10 66]<br>[-4 12 66] |
|  | Right angular gyrus | 143 | **2.50e-4**<br>**1.37e-6**<br>**0.028** | **0.027**<br>**0.008**<br>0.496 | F(2,101)=17.66<br><sup>a</sup>pre-ECT > no-depression<br><sup>a</sup>pre-ECT > non-ECT | [36 -56 28] |
| $Q_{s_t, a_t, chosen}^N$ | | | | | NS | |
| $Q_{s_t, a_t, unchosen}^N$ | | | | | NS | |
| **Subjective Experience Computations** |  |  |  |  |  |  |
| $\beta_{+\delta_t^P}$ | | | | | NS | |
| $\beta_{-\delta_t^P}$ | | | | | NS | |
| $\beta_{Q_{s_t, a_t, chosen}^P}$ | | | | | NS | |
| $\beta_{Q_{s_t, a_t, unchosen}^P}$ | | | | | NS | |
| $\beta_{+\delta_t^N}$ | Medial prefrontal cortex | 279 | **8.44e-7**<br>**3.43e-10** | 0.351<br>**0.048** | F(2,101)=13.42<br><sup>a</sup>no-depression > pre-ECT | [0 56 24] |
|  | Ventromedial prefrontal cortex | 74 | **0.020**<br>**1.63e-9** | 0.877<br>0.266 | F(2,101)=11.09<br><sup>a</sup>no-depression > pre-ECT | [0 58 -2] |
| $\beta_{-\delta_t^N}$ | Left supplementary motor area | 43 | 0.140<br>0.080 | **0.036**<br>**0.022** | F(2,101)=17.23<br><sup>a</sup>pre-ECT > no-depression | [-4 10 66] |
|  | Right angular gyrus | 85 | **0.008**<br>**1.31e-4** | 0.117<br>**0.021** | F(2,101)=15.39<br><sup>a</sup>pre-ECT > no-depression | [34 -58 28] |
| $\beta_{Q_{s_t, a_t, chosen}^N}$ | | | | | NS | |
| $\beta_{Q_{s_t, a_t, unchosen}^N}$ | | | | | NS | |

All analyses were performed at an uncorrected threshold of $p < 0.001$ in which reported results were selected using an FWE-corrected threshold of $p < 0.05$ at cluster and peak voxel levels. Bold font indicates significance. NS: not significant. <sup>a</sup>post-hoc t-test.

**Fig. 2.**
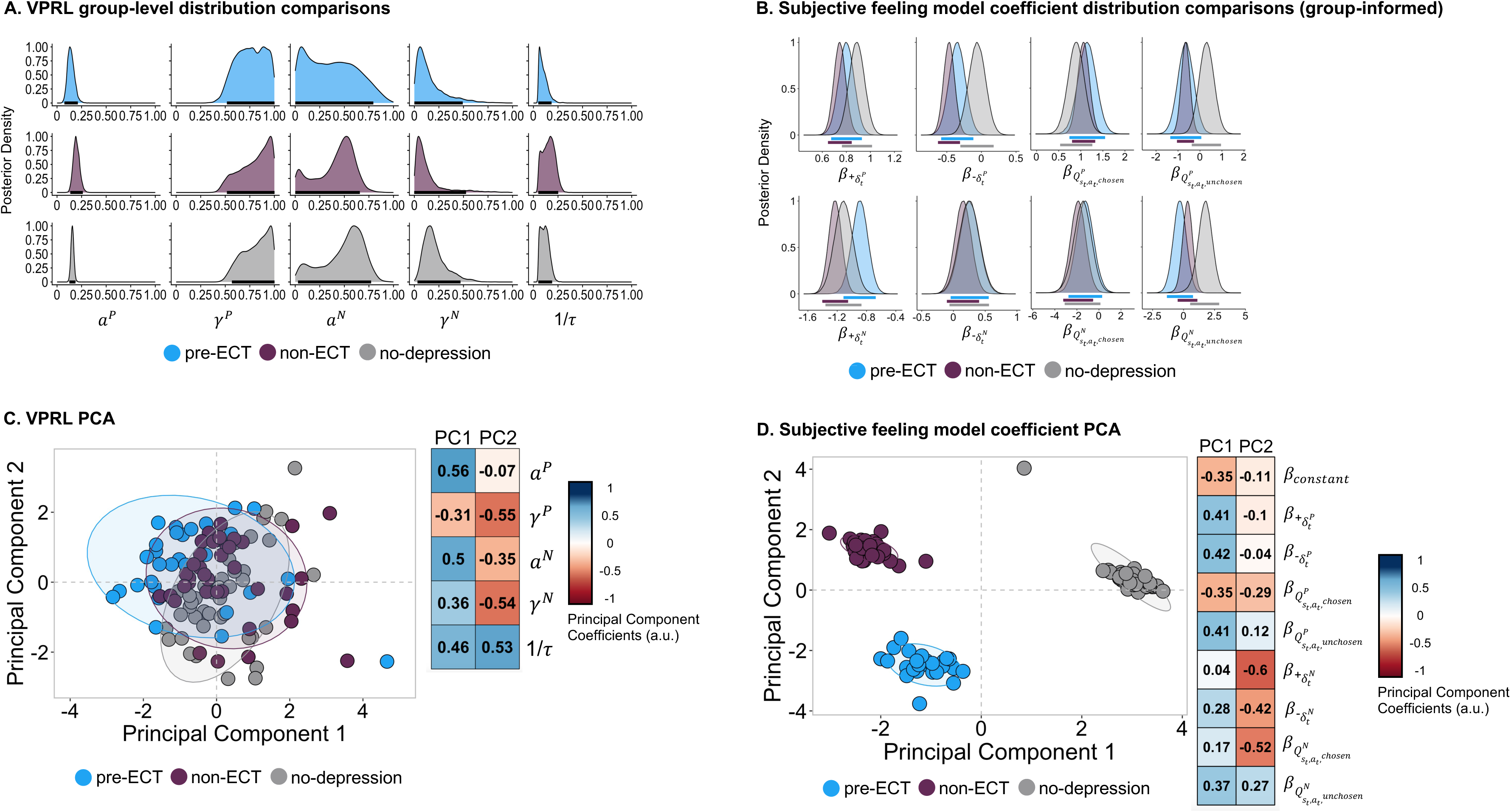
Mechanistic differences in learning and emotional phenotypes across major depression groups. Group-level posterior distributions of **(A)** valence-partitioned reinforcement learning (VPRL) model parameters and **(B)** coefficients from Subjective Feeling model that used participants’ expectations and prediction errors to predict subjective ratings during the task. Principal component (PC) analysis biplots with PC1 (x-axis) and PC2 (y-axis) illustrating cohort variability in **(C)** VPRL parameters and **(D)** Subjective Feeling model coefficients. Each point represents each participants’ individual-level median values from the group-informed models. PC 1 and 2 coefficients for respective variables are reported.

We observed group-level differences in the *positive system learning rate* (*α^P^*, Fig. 2A, eTable4). Non-ECT showed evidence of higher *α^P^* than both pre-ECT (median difference [95% HDI]=0.06[-0.04,0.15]) and no-depression (0.04[-0.03,0.11]). Upon visual inspection (Fig. 2A), the posterior distributions indicated large within-group variability for γ*^P^, a^N^*, and γ*^N^*. The median and 95% HDI for VPRL parameters (*α^P^, α^N^, γ^P^, γ^N^*, and τ) for each group and group-level comparisons are reported (eTable4).

We performed a principal components analysis to reduce dimensionality and assess whether distinct covariation amongst the five-vector of VPRL parameters could distinguish the three groups (Fig. 2C). The combination of the first two principal components (PCs) explained 63.2% variance and reveals a relationship between depression status and PC1 loading. Fig. 2C shows information about how each parameter contributed to PC1.

### Neural

We compared BOLD response differences associated with VPRL signals across the three groups (Table 1). We found significant differences in the BOLD response to the positive system Q-values 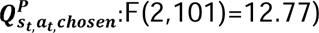 and negative system prediction errors 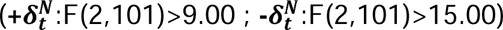 in the right calcarine sulcus, medial and ventromedial prefrontal cortex, left supplementary motor cortex, and right angular gyrus. Refer to Table 1 for details. eTable 5 describes brain areas tracking positive and negative system learning signals across participants.

### Behavioral and Neural *Computational Phenotypes* for Subjective Feelings in depression groups Behavioral

We hypothesized that VPRL learning signals may drive changes in subjective feelings about the consequences of choices made (Fig. 1B). We hypothesized a simple linear combination of these signals (and included a constant term to represent a baseline feeling state): 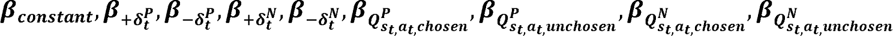 to predict subjective ratings. Group-level parameter estimates and comparisons of posterior distributions are shown in Fig. 2B and eTable6.

Both depression groups’ parameter estimates compared to the no-depression group) showed stronger influences of positive system prediction errors for less rewarding outcomes 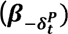 and counterfactual choice expectations 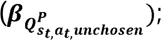 however, no-depression participants showed a stronger influence of negative system counterfactual choice expectations 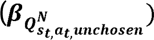 on feelings. Pre-ECT patients showed a stronger influence of negative system prediction errors for worse punishments 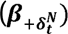 relative to non-ECT participants. No-depression participants showed a stronger influence of positive system prediction errors for more rewarding outcomes 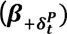 relative to non-ECT participants.

Group-level variation in nearly all Subjective Feeling model parameters (Fig. 2B) suggest high-dimensionality in how affective responses are driven across the three groups. Therefore, we performed PCA. The first two PCs explained 91.4% variance and revealed natural clustering of each group (Fig. 2D). Fig. 2D shows information about how parameters contribute to PC directions.

### Neural

We compared BOLD response differences associated with the influence of VPRL signals on subjective feelings across the three groups (Table 1). The medial and ventromedial prefrontal cortex, left supplementary motor area, and right angular gyrus showed significantly greater BOLD responses to negative system outcome prediction errors 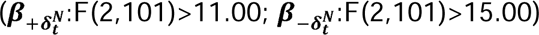 in no-depression versus pre-ECT groups. See Table 1 and eMethods for full statistical details.

### *Computational phenotype*s of dynamic learning and associated affective dynamics may be predictive of depression status

The preceding analyses used hierarchical Bayesian methods for model fitting that incorporated prior knowledge of the clinical status when fitting group-informed parameters (eTable3). We tested the hypothesis that VPRL and Subjective Feeling models, fit to behavioral dynamics on the PRPwSR task alone, would be sufficient to identify and discriminate patients with versus without depression. To test this, we fit model parameters for each participant without group information included at any stage. We then reduced the resulting 14-vector of parameters 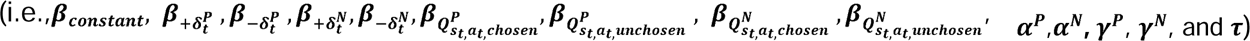 to two dimensions using PCA. While the first two PCs explain only 33.3% of the total variance, color-coding depression status clearly shows a pattern that distinguishes no-depression from positive-depression status (Fig. 3A). eFig.5 shows pairs plots to reveal the underlying data structure for all combinations of resulting PCs.

**Fig. 3.**
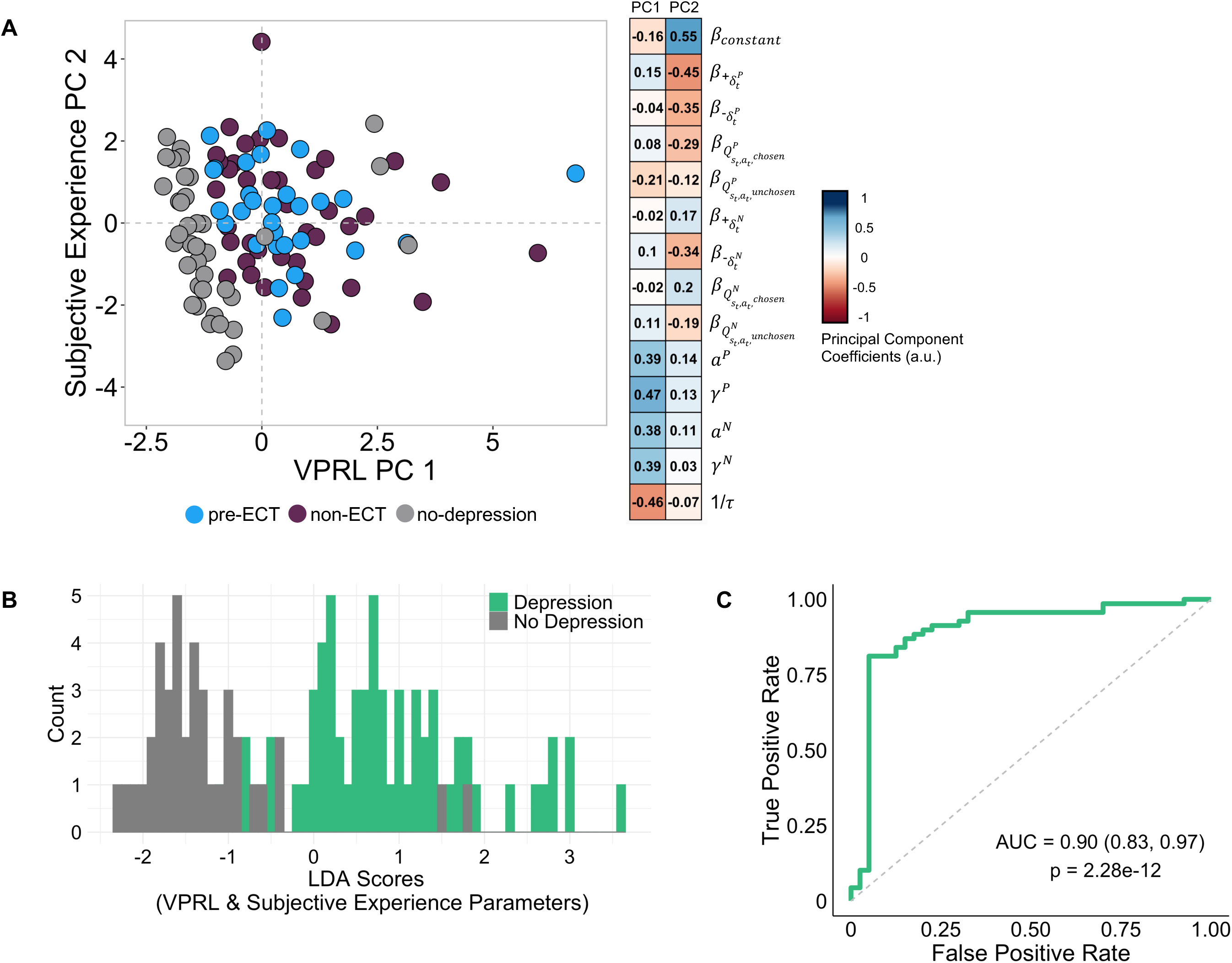
Leave-one-out cross-validation linear discriminant analysis of depression diagnosis. **(A)** Principal component (PC) analysis biplot of individually estimated (i.e., non-group informed) valence partitioned reinforcement learning (VPRL) parameters and Subjective Feeling model coefficients with ‘VPRL’ PC1 on the x-axis and ‘Subjective Experience’ PC2 on the y-axis (left). Heatmap of learning and affective parameters for the first two PCs where VPRL parameters described PC 1 and affective parameters described PC 2 (right). **(B)** Individual’s individual-level VPRL and affective parameters were inserted into a leave-one-out cross-validated linear discriminant analysis (LDA) to determine their predictive accuracy on depression diagnosis; distribution of participants’ resulting LDA scores by depression (green, N=69) and no depression (grey, N=40) groups. **(C)** Receiver Operating Characteristic curve showing comparison of true and false positive rates for depression versus no depression diagnosis in participants, with associated area under the curve (AUC).

We next sought to determine whether individually fit learning and subjective experience parameters could predict depression status. We performed a supervised, leave-one-out-cross-validated, linear discriminant analysis (LDA) using each individual’s 14-vector of parameters as the independent set of variables and their known clinical status as the dependent categorical variable. LDA score distributions are shown in Fig. 3B for depression and no depression groups. The overall prediction accuracy of the leave-one-out LDA model was 86% (p-value=3.08e-7, 95% CI=0.77, 0.91) with 0.85 sensitivity and 0.86 specificity. The resulting ROC curve (Fig. 3C) shows excellent diagnostic performance compared to chance with an AUC=0.90; 95%CI=[0.83-0.97], p-value=2.28e-12. See eTable7 for more information on model performance.

## DISCUSSION

We investigated neurobehavioral dynamics of learning and associated moment-to-moment changes in feelings in 1) patients with TRD planning to undergo ECT; 2) patients with depression managed without plans for ECT; and 3) participants without depression. We report observations from the first of two observational research visits (Fig. 1A). VPRL models of behavior on the PRPwSR task (Fig. 1B) yielded a set of parameters that suggested a spectrum of depression (Fig. 2C) and provided a means to quantitatively estimate moment-to-moment changes in subjective feelings (eFig.4). Notably, Subjective Feeling models further distinguished the three depression categories (Fig. 2D). In addition to these behavioral results, the derived *computational phenotypes* differentiated fMRI BOLD-response patterns associated with learning and affective dynamics (Table 1). Together, these results suggest that computational descriptions of learning and affective behavior may be sufficient to characterize patients with depression. Indeed, VPRL and Subjective Feeling model parameters fit to individuals’ behavior yielded an unbiased computational phenotype that provided excellent sensitivity and specificity for depression-state classification (Fig. 3).

Neurobehavioral dynamics associated with learning and subjective experience are complex and traditionally challenging to characterize. Computational psychiatric approaches provide a potential solution^59–61^. Prior work described neurocomputational processes associated with momentary happiness^13^ and changes in learning, dependent on whether the trial was rewarding or punishing, associated with depression^12^. The present study is, to our knowledge, the first to apply computational psychiatric methods to TRD and to combine insights from valence-partitioning RL methods to infer subjective experience.

The VPRL model we applied was recently shown to track sub-second changes in human dopamine levels^16^. Those data showed that dopamine reacted systematically to positive system prediction errors 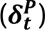 and negative system prediction errors 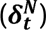^16^. In the present study, these signals differentially modulated learning and affective behavior in depression (Figs. 2-3). VPRL positive-system learning rates (*α^P^*) were decreased in pre-ECT patients but increased in non-ECT patients (Fig. 2A) whose symptoms were more effectively managed (eTables1-2). Further, negative-system learning rates (*α^N^*) appear to be decreased in both depression groups (Fig. 2A). In these models, learning rates relate how the magnitude of the error signals 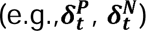 are weighed in updating expectations. Consistent with this conceptualization, we also observed that error signal weights on subjective feeling reports 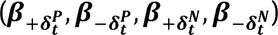 were significantly altered in patients with depression (Fig. 2B). BOLD-responses to 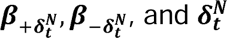, and also differed between pre-ECT and no-depression groups (Table 1). Together our results suggest that regions in the brain associated with dopaminergic prediction error processing may be altered in patients with depression, but particularly so in relation to negative-system processes in TRD.

Group-informed *computational phenotypes* (Fig. 2) provided insight into neurobehavioral dynamics that differentiated the groups in the present work. However, to determine whether computational phenotypes may inform more broadly, we simulated ignorance of clinical information by fitting models to individuals while excluding prior clinical knowledge (Fig. 3,eFig.5). This degraded the statistical quality of the models (eTable3) but allowed us to test the hypothesis that unbiased modeling may be used for diagnostic purposes. Indeed, PCA separation between the *three* groups was not as clear using individually-fit parameters (Fig. 3A) compared to group-fit parameters (Fig. 2D), but depression versus no-depression categories were still clear (Fig. 3A) and leave-one-out-cross-validated modeling yielded excellent classification performance (Fig. 3D). Together, these results support the notion that *computational phenotypes* based on hypothesis-driven models may be developed to augment diagnosis methods and form a basis for further research into neurobehavioral mechanisms underlying psychiatric conditions. However, we note that more work is needed before the results here can be used clinically.

## CONCLUSIONS

Subjective feelings are a critical element in our interpretation of sensory information and the choices we make as autonomous humans. The challenges in investigating subjective experience are its inherently private nature and the fact that observable behavior and latent conscious experiences do not always appear to be consistent. This represents a major barrier for psychiatric medicine and consciousness research. In this study, we used computational models grounded in RL theory and prior empirical work in computational neuroscience to summarize the relationship between observable objective stimuli, choice behaviors, brain responses, and unobservable but behaviorally inferred subjective feelings. Depression, including TRD, is at its core a disease of private experience that can manifest as sometimes obvious but often subtle changes in behavior. Using a relatively simple task that requires adaptive changes in behavior but also intermittently recorded subjective self-reported experiences, we characterized differences in neurobehavioral signals in patients with TRD, but also depression versus no-depression more generally. Future work will characterize neurobehavioral changes following ECT (i.e.,“visit 2”,Fig. 1A)^62^.

## Supporting information

Supplimental Content

## Data Availability

All data produced in the present study are available upon reasonable request to

## ACKNOWLEDGEMENTS

None.

## Author Contributions

Drs. Kishida and Jones had full access to all the data in the study and take responsibility for the integrity of the data and the accuracy of the data analysis.

*Study concept and design:* Gligorovic, Jones, Kishida. *Acquisition, analysis, or interpretation of data:* All authors. *Drafting of the manuscript:* Jones, Kishida.

*Critical revision of the manuscript for important intellectual content:* All authors.

*Statistical analysis:* Jones, Sands, Trattner.

*Obtained funding:* Sands and Kishida.

*Administrative, technical, or material support:* All authors.

*Study supervision:* Gligorovic, Douglas, Ramos, Jones, Kishida.

## Additional Contributions

We thank all of the individuals who took part in this study, clinic staff who assisted with recruitment, and MRI staff who assisted with research visits. We thank Dr. Mary Moya-Mendez for her work in community advertisement and IRB development. The following research assistants from the Translational Neuroscience Department at Wake Forest School of Medicine were involved in data collection: Ashley Shipp, BS and Michael Howze, BS.

## REFERENCES

1. World Health Organization. The global burden of disease: 2004 update. Published online 2008:146. Accessed June 28, 2023. https://apps.who.int/iris/handle/10665/43942

2. Akil H, Gordon J, Hen R, et al. Treatment resistant depression: A multi-scale, systems biology approach. Neurosci Biobehav Rev. 2018;84:272–288. doi:10.1016/j.neubiorev.2017.08.019

3. Gaynes BN, Lux L, Gartlehner G, et al. Defining treatment-resistant depression. Depress Anxiety. 2020;37(2):134-145. doi:10.1002/da.22968

4. Warden D, Rush AJ, Trivedi MH, Fava M, Wisniewski SR. The STAR*D project results: A comprehensive review of findings. Curr Psychiatry Rep. 2007;9(6):449–459. doi:10.1007/s11920-007-0061-3

5. Magid M, Kellner CH, Greenberg RM. ISEN:: What is ECT. International Society for ECT and Neurostimulation. Accessed June 28, 2023. https://www.isen-ect.org/what-is-ect

6. Huys QJM, Daw ND, Dayan P. Depression: A Decision-Theoretic Analysis. Annu Rev Neurosci. 2015;38(1):1-23. doi:10.1146/annurev-neuro-071714-033928

7. Williams LM. Precision psychiatry: a neural circuit taxonomy for depression and anxiety. Lancet Psychiatry. 2016;3(5):472–480. doi:10.1016/S2215-0366(15)00579-9

8. Berwian IM, Wenzel JG, Collins AGE, et al. Computational Mechanisms of Effort and Reward Decisions in Patients With Depression and Their Association With Relapse After Antidepressant Discontinuation. JAMA Psychiatry. 2020;77(5):513–522. doi:10.1001/jamapsychiatry.2019.4971

9. Chen C, Takahashi T, Nakagawa S, Inoue T, Kusumi I. Reinforcement learning in depression: A review of computational research. Neurosci Biobehav Rev. 2015;55:247–267. doi:10.1016/j.neubiorev.2015.05.005

10. Adams RA, Huys QJM, Roiser JP. Computational Psychiatry: towards a mathematically informed understanding of mental illness. J Neurol Neurosurg Psychiatry. 2016;87(1):53–63. doi:10.1136/jnnp-2015-310737

11. Robinson OJ, Chase HW. Learning and Choice in Mood Disorders: Searching for the Computational Parameters of Anhedonia. Comput Psychiatry Camb Mass. 2017;1:208–233. doi:10.1162/CPSY_a_00009

12. Brown VM, Zhu L, Solway A, et al. Reinforcement Learning Disruptions in Individuals With Depression and Sensitivity to Symptom Change Following Cognitive Behavioral Therapy. JAMA Psychiatry. 2021;78(10):1–11. doi:10.1001/jamapsychiatry.2021.1844

13. Rutledge RB, Moutoussis M, Smittenaar P, et al. Association of Neural and Emotional Impacts of Reward Prediction Errors With Major Depression. JAMA Psychiatry. 2017;74(8):790–797. doi:10.1001/jamapsychiatry.2017.1713

14. Rothkirch M, Tonn J, Köhler S, Sterzer P. Neural mechanisms of reinforcement learning in unmedicated patients with major depressive disorder. Brain. 2017;140(4):1147–1157. doi:10.1093/brain/awx025

15. Kishida KT, Sands LP. A Dynamic Affective Core to Bind the Contents, Context, and Value of Conscious Experience. In: Waugh CE, Kuppens P, eds. Affect Dynamics. Springer International Publishing; 2021:293–328. doi:10.1007/978-3-030-82965-0_12

16. Sands LP, Jiang A, Liebenow B, et al. Subsecond fluctuations in extracellular dopamine encode reward and punishment prediction errors in humans. Sci Adv. 9(48):eadi4927. doi:10.1126/sciadv.adi4927

17. Sands LP, Jiang A, Jones RE, Trattner JD, Kishida KT. Valence-partitioned learning signals drive choice behavior and phenomenal subjective experience in humans. Published online March 18, 2023: 2023.03.17.533213. doi:10.1101/2023.03.17.533213

18. Insel T, Cuthbert B, Garvey M, et al. Research Domain Criteria (RDoC): Toward a New Classification Framework for Research on Mental Disorders. Am J Psychiatry. 2010;167(7):748–751. doi:10.1176/appi.ajp.2010.09091379

19. Taylor CT, Pearlstein SL, Stein MB. A tale of two systems: Testing a positive and negative valence systems framework to understand social disconnection across anxiety and depressive disorders. J Affect Disord. 2020;266:207–214. doi:10.1016/j.jad.2020.01.041

20. Groenewold NA, Opmeer EM, de Jonge P, Aleman A, Costafreda SG. Emotional valence modulates brain functional abnormalities in depression: Evidence from a meta-analysis of fMRI studies. Neurosci Biobehav Rev. 2013;37(2):152–163. doi:10.1016/j.neubiorev.2012.11.015

21. Medeiros GC, Rush AJ, Jha M, et al. Positive and negative valence systems in major depression have distinct clinical features, response to antidepressants, and relationships with immunomarkers. Depress Anxiety. 2020;37(8):771–783. doi:10.1002/da.23006

22. Hill KE, Pegg S, Dao A, et al. Characterizing positive and negative valence systems function in adolescent depression: An RDoC-informed approach integrating multiple neural measures. J Mood Anxiety Disord. 2023;3:100025. doi:10.1016/j.xjmad.2023.100025

23. Paulus MP, Stein MB, Craske MG, et al. Latent variable analysis of positive and negative valence processing focused on symptom and behavioral units of analysis in mood and anxiety disorders. J Affect Disord. 2017;216:17–29. doi:10.1016/j.jad.2016.12.046

24. American Psychiatric Association, American Psychiatric Association, eds. Diagnostic and Statistical Manual of Mental Disorders: DSM-5. 5th ed. American Psychiatric Association; 2013.

25. Montague PR, Dayan P, Sejnowski TJ. A framework for mesencephalic dopamine systems based on predictive Hebbian learning. J Neurosci. 1996;16(5):1936–1947. doi:10.1523/JNEUROSCI.16-05-01936.1996

26. Schultz W, Dayan P, Montague PR. A Neural Substrate of Prediction and Reward. Science. 1997;275(5306):1593-1599. doi:10.1126/science.275.5306.1593

27. Kumar P, Waiter G, Ahearn T, Milders M, Reid I, Steele JD. Abnormal temporal difference reward-learning signals in major depression. Brain. 2008;131(8):2084–2093. doi:10.1093/brain/awn136

28. Rupprechter S, Romaniuk L, Series P, et al. Blunted medial prefrontal cortico-limbic reward-related effective connectivity and depression. Brain. 2020;143(6):1946–1956. doi:10.1093/brain/awaa106

29. Bakic J, Pourtois G, Jepma M, Duprat R, De Raedt R, Baeken C. Spared internal but impaired external reward prediction error signals in major depressive disorder during reinforcement learning. Depress Anxiety. 2017;34(1):89–96. doi:10.1002/da.22576

30. Rutledge RB, Skandali N, Dayan P, Dolan RJ. A computational and neural model of momentary subjective well-being. Proc Natl Acad Sci. 2014;111(33):12252–12257. doi:10.1073/pnas.1407535111

31. Eldar E, Niv Y. Interaction between emotional state and learning underlies mood instability. Nat Commun. 2015;6(1):6149. doi:10.1038/ncomms7149

32. Huys QJM, Renz D. A Formal Valuation Framework for Emotions and Their Control. Biol Psychiatry. 2017;82(6):413–420. doi:10.1016/j.biopsych.2017.07.003

33. Katahira K, Fujimura T, Okanoya K, Okada M. Decision-Making Based on Emotional Images. Front Psychol. 2011;2. doi:10.3389/fpsyg.2011.00311

34. Etkin A, Büchel C, Gross JJ. The neural bases of emotion regulation. Nat Rev Neurosci. 2015;16(11):693–700. doi:10.1038/nrn4044

35. Liebenow B, Jones R, DiMarco E, et al. Computational reinforcement learning, reward (and punishment), and dopamine in psychiatric disorders. Front Psychiatry. 2022;13. Accessed October 20, 2022. https://www.frontiersin.org/articles/10.3389/fpsyt.2022.886297

36. Chase HW, Frank MJ, Michael A, Bullmore ET, Sahakian BJ, Robbins TW. Approach and avoidance learning in patients with major depression and healthy controls: relation to anhedonia. Psychol Med. 2010;40(3):433–440. doi:10.1017/S0033291709990468

37. Dombrovski AY, Szanto K, Clark L, et al. Corticostriatothalamic reward prediction error signals and executive control in late-life depression. Psychol Med. 2015;45(7):1413–1424. doi:10.1017/S0033291714002517

38. Dombrovski AY, Szanto K, Clark L, Reynolds CF III, Siegle GJ. Reward Signals, Attempted Suicide, and Impulsivity in Late-Life Depression. JAMA Psychiatry. 2013;70(10):1020–1030. doi:10.1001/jamapsychiatry.2013.75

39. Lawson RP, Nord CL, Seymour B, et al. Disrupted habenula function in major depression. Mol Psychiatry. 2017;22(2):202–208. doi:10.1038/mp.2016.81

40. Liu WH, Valton V, Wang LZ, Zhu YH, Roiser JP. Association between habenula dysfunction and motivational symptoms in unmedicated major depressive disorder. Soc Cogn Affect Neurosci. 2017;12(9):1520–1533. doi:10.1093/scan/nsx074

41. Rolls ET. A non-reward attractor theory of depression. Neurosci Biobehav Rev. 2016;68:47–58. doi:10.1016/j.neubiorev.2016.05.007

42. McCabe C, Woffindale C, Harmer CJ, Cowen PJ. Neural Processing of Reward and Punishment in Young People at Increased Familial Risk of Depression. Biol Psychiatry. 2012;72(7):588–594. doi:10.1016/j.biopsych.2012.04.034

43. Dayan P, Niv Y. Reinforcement learning: The Good, The Bad and The Ugly. Curr Opin Neurobiol. 2008;18(2):185–196. doi:10.1016/j.conb.2008.08.003

44. Pessiglione M, Delgado MR. The good, the bad and the brain: neural correlates of appetitive and aversive values underlying decision making. Curr Opin Behav Sci. 2015;5:78–84. doi:10.1016/j.cobeha.2015.08.006

45. Sutton RS, Barto AG. Introduction to Reinforcement Learning. Vol 135. MIT press Cambridge; 1998.

46. Collins AGE, Frank MJ. Opponent actor learning (OpAL): Modeling interactive effects of striatal dopamine on reinforcement learning and choice incentive. Psychol Rev. 2014;121(3):337–366. doi:10.1037/a0037015

47. Daw ND, Kakade S, Dayan P. Opponent interactions between serotonin and dopamine. Neural Netw. 2002;15(4):603–616. doi:10.1016/S0893-6080(02)00052-7

48. Huys QJM, Cools R, Gölzer M, et al. Disentangling the Roles of Approach, Activation and Valence in Instrumental and Pavlovian Responding. PLOS Comput Biol. 2011;7(4):e1002028. doi:10.1371/journal.pcbi.1002028

49. Spitzer KK, Williams JBW. Patient Health Questionnaire-9. APA PsycTests. 10.1037/t06165-000

50. Hamilton M. A RATING SCALE FOR DEPRESSION. J Neurol Neurosurg Psychiatry. 1960;23(1):56–62. Accessed June 28, 2023. https://www.ncbi.nlm.nih.gov/pmc/articles/PMC495331/

51. Nasreddine ZS, Phillips NA, Bédirian V, et al. The Montreal Cognitive Assessment, MoCA: A Brief Screening Tool For Mild Cognitive Impairment. J Am Geriatr Soc. 2005;53(4):695–699. doi:10.1111/j.1532-5415.2005.53221.x

52. Kruschke J. Doing Bayesian Data Analysis: A Tutorial with R, JAGS, and Stan. Academic Press; 2014.

53. Kishida KT, Montague PR. Economic probes of mental function and the extraction of computational phenotypes. J Econ Behav Organ. 2013;94:234–241. doi:10.1016/j.jebo.2013.07.009

54. Schurr R, Reznik D, Hillman H, Bhui R, Gershman SJ. Dynamic computational phenotyping of human cognition. Nat Hum Behav. 2024;8(5):917–931. doi:10.1038/s41562-024-01814-x

55. Sutton RS, Barto AG. Reinforcement Learning: An Introduction. MIT press; 2018.

56. Carpenter B, Gelman A, Hoffman MD, et al. Stan: A Probabilistic Programming Language. J Stat Softw. 2017;76:1. doi:10.18637/jss.v076.i01

57. Ahn WY, Haines N, Zhang L. Revealing Neurocomputational Mechanisms of Reinforcement Learning and Decision-Making With the hBayesDM Package. Comput Psychiatry Camb Mass. 2017;1:24–57. doi:10.1162/CPSY_a_00002

58. Kruschke JK. What to believe: Bayesian methods for data analysis. Trends Cogn Sci. 2010;14(7):293–300. doi:10.1016/j.tics.2010.05.001

59. Montague PR, Dolan RJ, Friston KJ, Dayan P. Computational psychiatry. Trends Cogn Sci. 2012;16(1):72-80. doi:10.1016/j.tics.2011.11.018

60. Wang XJ, Krystal JH. Computational Psychiatry. Neuron. 2014;84(3):638-654. doi:10.1016/j.neuron.2014.10.018

61. Huys QJM, Maia TV, Frank MJ. Computational psychiatry as a bridge from neuroscience to clinical applications. Nat Neurosci. 2016;19(3):404–413. doi:10.1038/nn.4238

62. Jones R, Sands LP, Trattner JD, et al. 153. Electroconvulsive Therapy Changes the Emotional Impact of Learning Signals in Patients With Treatment-Resistant Depression. Biol Psychiatry. 2024;95(10):S161–S162. doi:10.1016/j.biopsych.2024.02.388

