## Supplementary material for "Computational Phenotyping of Treatment-Resistant Depression prior to Electroconvulsive Therapy": Supplimental Content: Final visit 1 - Supplementary Online Content (KTK Approved FINAL).docx

**eMethods.**

**eReferences.**

**eFigure 1.** Probabilistic Reward and Punishment with Subjective Rating (PRPwSR) task phases and icon values

**eFigure 2.** Expected versus actual icon value for each cohort

**eFigure 3.** PRPwSR task performance metrics by cohort

**eFigure 4.** Model-predicted versus participant-reported ratings

**eFigure 5.** Pairs plots of individually-fit learning parameter and subjective rating coefficient PCA

**eTable 1.** Participant demographics and clinical characteristics

**eTable 2.** Current psychiatric medications and comorbidities reported by participants with depression

**eTable 3.** Performance metrics of different group-level prior distributions

**eTable 4.** Comparison of VPRL model parameter posterior distributions across cohorts

**eTable 5.** Positive and negative system prediction error and expected value results from whole-brain one-sample t-test pooling all participants

**eTable 6.** Comparison of Subjective Feeling model coefficient posterior distributions across cohorts

**eTable 7.** Predicted versus actual depressions tatus from leave-one-out cross-validation linear discriminant analysis

**eMethods**

**Study Design**

This study involved two research visits that followed the standard of care ECT treatment timeline (Fig. 1A) — here, we focus on data collected during the “pre-ECT” research “visit 1”. We recruited three participant groups according to depression diagnosis and ECT treatment plan (pre-ECT depression, non-ECT depression, and no-depression groups). All participants completed the Probabilistic Reward and Punishment with Subjective Rating (PRPwSR) task^1,2^ with fMRI scanning on both visit 1 and visit 2. Clinical characterization (including the Patient Health Questionnaire 9 (PHQ-9^3^), Hamilton Depression Rating Scale (HAM-D^4^), and Montreal Cognitive Assessment (MOCA^5^)) was performed after fMRI scanning was completed on each visit. Visit 1 and visit 2 were separated by the duration it took for pre-ECT patients to complete their standard of care ECT series. pre-ECT patients then completed their second research visit 2 after their ECT series was deemed completed by their psychiatrist. No-depression and non-ECT depression participants did not undergo ECT treatment during the study period but completed their first and second research visits following a timeline matched to the ECT group. Thus, no-depression, non-ECT depression, and pre-ECT participants’ second research visit occurred approximately one to two month after research visit 1.

**Participants**

All participants in this study provided written informed consent and the protocol was approved by the Institutional Review Board (IRB00056131) of Wake Forest University School of Medicine. For all participants, inclusion criteria included ages 18 to 85 and English speaking while exclusion criteria included individuals with contraindications for MRI scanning, individuals not able to provide written consent and verbal assent, individuals not able to understand task instructions or consented documents, and women who are pregnant.

We recruited three cohorts of participants. Pre-ECT patients included patients with treatment-resistant depression naïve to ECT who consented to begin ECT treatment during a non-research standard of care visit at Atrium Health Wake Forest Baptist Outpatient Psychiatry and Behavioral Health clinic (pre-ECT cohort N=29; 55.17% female). Participants with (non-ECT N=40; 70% female) and without depression (no-depression N=41; 65.90% female) were recruited through advertisements in the local Winston-Salem, North Carolina community. Participants with depression reported a current diagnosis of depression on the study’s screening form or, after assessment in this study, scored mild, moderate, or greater levels of depression according to PHQ-9 and HAM-D assessments. These participants reported no prior history of ECT and did not plan to undergo ECT during the study period. Participants without depression consisted of community volunteers who reported no current diagnosis of depression and scored below threshold levels of depression severity on PHQ-9 and HAM-D assessments. During the analysis stage of our study, we re-assigned two individuals (from the no-depression recruitment group) to the non-ECT depression group; these participants had initially self-reported no diagnosis of depression, but upon assessment with both PHQ-9 and HAM-D measures they scored in the mild to moderate depression severity range.

Approximately half (n = 22) of non-ECT depression participants indicated trying two or more different medications in the past to treat their depression, meeting some criteria for treatment-resistant depression. However, participants in the non-ECT depression group indicated never receiving ECT in the past and did not receive ECT during the study period.

Two participants with depression and three no-depression participants were excluded in study analyses due to missing game data. One no-depression participant was excluded from individual-level Subjective Feeling model fitting and eFig.4 analyses due to reporting the same rating for all trials. eTable1 contains information about participant demographic and clinical characteristics and eTable2 describes current psychiatric medications and comorbidities reported by depression cohorts.

**Probabilistic Reward and Punishment with Subjective Rating (PRPwSR) Task**

The PRPwSR task is a 150-trial, two-alternative forced choice task (Fig.1B). The tasks transitions through three phases with PI: 25 trials, PII: 50 trials, and PIII: 75 trials. In PI participants learn about three cues that lead to a $1 reward 25%, 50%, or 75% of the time. In PII they learn (on half of the 50 trials) about three new cues that lead to a $1 loss 25%, 50%, or 75% of the time; on the other half of trials in PII they continue to learn about the three reward cues. In PIII, participants experience trials were two of any of the six cues may be presented together and the probability of outcome is the same, but the magnitude of reward or punishment is shifted (see eFig.1) so that the ‘best choice’ must be re-learned.

After each trial throughout the task, there is a 1/3 probability that a rating screen would be presented after the outcome for that trial was shown. The rating screen queried participants, “How do you feel about the last outcome?” Participants reported their subjective feelings on a scale ranging from “very bad” to “very good.” See eFig.1 for additional information regarding the PRPwSR task structure and eFig.2 and eFig.3 for information about participant performance on the PRPwSR task.

**Computational Modeling**

We used hierarchical Bayesian analysis^6^ to fit the valence-partitioned reinforcement learning (VPRL) computational model^1,2,7^ to participants’ decision-making behavior on the PRPwSR task (Fig.2). Choice behavior was modeled strictly as a learning problem in a manner consistent with standard RL methods. Expected values and outcome prediction errors derived specifically from the VPRL framework were then used to fit a linear regression model to predict subjective ratings^2^ on those trials where participants were asked ‘how they felt’ about the outcome they had just experienced. Each individual was then assumed to be characterized by their unique set of behaviorally derived parameters, which we refer to as their *computational phenotype*^8^.

*“Valence Partitioned Reinforcement Learning” Model of Choice Behavior*

The VPRL framework describes the specific hypotheses that independent positive- and negative-valence systems track expectations about rewarding and/or punishing contexts^1,2,7^. This approach was previously demonstrated to be a better fit to behavior on the PRPwSR task (compared to traditional univalent temporal difference RL models) and was shown to better explain sub-second fluctuations in extracellular dopamine levels in humans performing the PRPwSR task.^1^

In VPRL, temporal difference based Q-learning and prediction error models are used in parallel for the positive system (Eq. 1 and 2) and negative system (Eq. 3 and 4).

$$Q_{s_{t,}a_{t}}^{P}\leftarrow Q_{s_{t,}a_{t}}^{P}+\alpha^{P}\cdot\delta_{t}^{P}$$

*(1)*

$$\delta_{t}^{P}=\left\{ \begin{aligned} {outcome}_{t}+\gamma^{P}maxQ_{s_{t+1,}\tilde{\alpha}}^{P}-Q_{s_{t,}a_{t}}^{P} if {outcome}_{t}>0 \\ 0 +\gamma^{P}maxQ_{s_{t+1,}\tilde{\alpha}}^{P}-Q_{s_{t,}a_{t}}^{P} if {outcome}_{t}\leq0 \end{aligned} \right.$$

*(2)*

$$Q_{s_{t,}a_{t}}^{N}\leftarrow Q_{s_{t,}a_{t}}^{N}+\alpha^{N}\cdot\delta_{t}^{N}$$

*(3)*

$$\delta_{t}^{N}=\left\{ \begin{aligned} {|outcome}_{t}|+\gamma^{N}maxQ_{s_{t+1,}\tilde{\alpha}}^{N}-Q_{s_{t,}a_{t}}^{N} if {outcome}_{t}<0 \\ 0 +\gamma^{N}maxQ_{s_{t+1,}\tilde{\alpha}}^{N}-Q_{s_{t,}a_{t}}^{N} if {outcome}_{t}\geq0 \end{aligned} \right.$$

*(4)*

The superscripts *P* and *N* are used to denote the positive and negative systems, respectively. In Eq. 1, the positive system’s value estimate $Q_{s_{t,}a_{t}}^{P}$, of the quality, $Q$, of an action, $a_{t}$, in a given state, $s_{t}$, at time, $t$, is updated by the positive system’s reward prediction error, $\delta_{t}^{P}$. “$\alpha^{P}$” is the positive system’s learning rate that weights the influence of $\delta_{t}^{P}$updates of the value estimate, $Q_{s_{t,}a_{t}}^{P}.$In Eq. 2, ${outcome}_{t}$ is the reward experienced at time *t*. The discounting factor ($\gamma^{P}$) weighs the extent that the value of the future is considered; $maxQ_{s_{t+1,}\tilde{\alpha}}^{P}$ represents the value of the future state after choosing action $\alpha$ from the available actions ($\tilde{\alpha}$) that maximize the expected value ($maxQ^{P}$) in the future state ($s_{t+1}$). Eq. 3 and 4 describe the negative system’s value estimates ($Q_{s_{t,}a_{t}}^{N}$) and punishment prediction errors ($\delta_{t}^{N}$).

The overall estimate of value is obtained by integrating the independently estimated positive and negative system Q-values (Eq. 5):

$$Q_{s_{t},a_{t}}= Q_{s_{t,}a_{t}}^{P}- Q_{s_{t,}a_{t}}^{N}$$

*(5)*

Participants’ overall estimated Q-values were inserted into a softmax policy function (Eq. 6) to estimate the probabilities of choosing icons when presented in the PRPwSR task:

$$P\left( choice={choice}_{1} \right| Q_{s_{t},{choice}_{1}}, Q_{s_{t},{choice}_{2}})= \frac{e^{Q_{s_{t},{choice}_{1}}/\tau}}{e^{Q_{s_{t},{choice}_{1}}/\tau}+ e^{Q_{s_{t},{choice}_{2}}/\tau}}$$

*(6)*

Here, “$\tau$” is the choice temperature parameter constrained to a range of 0-20 that captures how deterministically ($\tau$ closer to 0) versus randomly ($\tau$ closer to 1) participants distribute their choices given their current estimates of value.

For each of the three cohorts (pre-ECT, non-ECT, no-depression), we fit the same VRPL model using hierarchical Bayesian analysis, assuming uniform priors, to derive parameter estimates pooled toward the respective group mean^6^. See Sands et. al for a detailed description of the hierarchical Bayesian procedure^1,2^.

We ran Hamiltonian Monte Carlo with No-U-Turn Sampler (HMC-NUTS) via Stan’s rstan interface to efficiently estimate the posterior distributions of VPRL model parameters across cohort data (version 4.2.2)^9^. For each cohort model fit, we ran four Markov chains, with 12,000 total samples per chain (8,000 after discarding warm-up samples). In addition, we confirmed that all Gelman-Rubin $\hat{R}$ values were approximately 1 for all parameters which indicated good chain mixing.

*“Subjective Feeling” Model of Emotional Experience to Choice Outcomes*

We next used objective learning signals, estimated within the VPRL framework, to determine whether these learning signals influenced participant-reported feelings on the PRPwSR task. For each subjective rating trial, we hypothesized that the subjective rating could be predicted by a linear combination of Q-values and outcome prediction errors for both the positive and negative valence systems. We also hypothesized that participants’ Q-values and prediction errors would differentially influence self-reported feelings in patients with TRD. Therefore, we modeled a linear regression with the subjective rating as the dependent variable and the Q-values ($\boldsymbol{Q}_{\boldsymbol{s}_{\boldsymbol{t,}}\boldsymbol{a}_{\boldsymbol{t,}}\boldsymbol{chosen}}^{\boldsymbol{P}}\boldsymbol{,}\boldsymbol{Q}_{\boldsymbol{s}_{\boldsymbol{t,}}\boldsymbol{a}_{\boldsymbol{t,}}\boldsymbol{unchosen}}^{\boldsymbol{P}}\boldsymbol{,}\boldsymbol{Q}_{\boldsymbol{s}_{\boldsymbol{t,}}\boldsymbol{a}_{\boldsymbol{t,}}\boldsymbol{chosen}}^{\boldsymbol{N}}\boldsymbol{,}\boldsymbol{Q}_{\boldsymbol{s}_{\boldsymbol{t,}}\boldsymbol{a}_{\boldsymbol{t,}}\boldsymbol{unchosen}}^{\boldsymbol{N}})$and outcome prediction errors (**+**$\boldsymbol{\delta}_{\boldsymbol{t}}^{\boldsymbol{P}}$, **-**$\boldsymbol{\delta}_{\boldsymbol{t}}^{\boldsymbol{P}}$, **+**$\boldsymbol{\delta}_{\boldsymbol{t}}^{\boldsymbol{N}}$, **-**$\boldsymbol{\delta}_{\boldsymbol{t}}^{\boldsymbol{N}}$) as the independent variables. The positive and negative sign on these error signals indicated greater-than (+) or less-than (-) expected and are separated for these models to reflect the hypothesis that these are not symmetrically weighted in their influence on affective responses. This linear regression model was fit using Bayesian methods^10^. We fit the regression model to each cohort (i.e., pre-ECT, non-ECT, no-depression) and implemented a leave-one-out cross-validation approach where each left-out participant’s posterior distributions of model coefficients were informed by their respective cohort. The Bayesian regression model generated posterior distributions of coefficient values for each of the eight independent learning variables (Fig.2B).

*Bayesian Subjective Feeling regression model*

We used (non-hierarchical) Bayesian linear regression^10^ to estimate participant’s momentary feelings ratings throughout the PRPwSR task using as predictors VPRL-model-derived expected values of chosen and unchosen options and prediction errors experienced during the 50 rating trials:

$$E\left( D_{i} | \beta,X \right)=\beta_{0}+\beta_{1}x_{1}+\beta_{2}x_{2}+\beta_{3}x_{3}+\beta_{4}x_{4}+\beta_{5}x_{5}+\beta_{6}x_{6}+\beta_{7}x_{7}+\beta_{8}x_{8}+\varepsilon_{i}$$

*(7)*

$$\varepsilon\sim Normal(0,\sigma^{2})$$

The expected value of a rating on trial *i* is denoted $E(D_{i}|\beta,X)$ and is assumed to be normally distributed. The mean of the subjective rating on a trial ($D_{i}$) is a linear function of positive and negative system expected value and prediction error predictor matrices ($X)$, with coefficients of the linear combination represented by $\beta$ vector. The predictor variables include Q-values of chosen and unchosen reward and loss icons ($Q_{s_{t,}a_{t,}chosen}^{P}, Q_{s_{t,}a_{t,}unchosen}^{P}, Q_{s_{t,}a_{t,}chosen}^{N}, Q_{s_{t,}a_{t,}unchosen}^{N})$ and reward and punishment prediction errors (+$\delta_{t}^{P}$, -$\delta_{t}^{P}$, +$\delta_{t}^{N}$, -$\delta_{t}^{N}$) that occur during the 50 rating trials for each participant, totaling eight model predictors. $\varepsilon_{i}$ are the normally-distributed errors with variance $\sigma^{2}.$ See Sands et. al for a detailed description of the Bayesian methods for the Subjective Feeling regression model^2^.

After excluding 1,000 warm-up samples, cohort model fits produced four parallel chains of length 2,500 for a total of 10,000 samples for each model parameter. $\hat{R}$ values were approximately 1 for all parameters.

**Functional MRI Analyses**

*Data acquisition*

For all participants, we acquired fMRI BOLD data by means of a multi-band echo-planar imaging (EPI) sequence using a Siemens MAGNETOM 3T Skyra whole-body scanner with a 32-channel head coil (MB factor = 8; TR = 1000ms; TE = 30ms; flip angle = 52 degrees; FOV = 20.8 cm; 72 interleaved sagittal slices; isotropic 2mm3 voxels) in addition to a high-resolution T1-weighted anatomical scan.

*Pre-processing*

We performed all data pre-processing using FSL^11^ and SPM12^12^. This included first estimating head motion via registration to a single-band reference image (SBRef); correcting for EPI (B0) distortion via a fieldmap estimated using reverse-phase encoded functional volumes (R-L and L-R directions) and FSL’s topup tool^13^; co-registering to the high-resolution (0.5x0.5x1 mm^3^) T1-weighted structural image and then to MNI template space; spatially smoothing with a 4mm FWHM Gaussian filter; high-pass filtering at 128sec (<0.008Hz); and normalizing by the session grand-mean value.

*Model-based analyses*

VPRL and subjective experience model terms were fit to each participant as described above. These terms were used to perform a fixed-effects analysis at the individual level. Contrasts generated at the individual level were then used to perform random effects analyses at the second level.

For each participant, we constructed two first-level general linear models (GLMs) to model blood-oxygen-level-dependent (BOLD) signals related to PRPwSR task events. Regressors of interest were convolved with a canonical hemodynamic response function. We assessed learning signals in the first GLM where regressors of interest included the option presentation timepoint parametrically modulated by participants’ expected values ($Q_{s_{t,}a_{t,}chosen}^{P}, Q_{s_{t,}a_{t,}unchosen}^{P}, Q_{s_{t,}a_{t,}chosen}^{N}, Q_{s_{t,}a_{t,}unchosen}^{N})$and the outcome presentation timepoint parametrically modulated by participants’ prediction errors (+$\delta_{t}^{P}$, -$\delta_{t}^{P}$, +$\delta_{t}^{N}$, -$\delta_{t}^{N}$) across all task trials. We performed an additional first-level GLM for each participant to model BOLD signals related to affective dynamics. Regressors of interest included the outcome presentation timepoint parametrically modulated by expected values and prediction errors weighted by respective Subjective Feeling model coefficient values: $Q_{s_{t,}a_{t,}chosen}^{P}\cdot\beta_{Q_{s_{t,}a_{t,}chosen}^{P}}, Q_{s_{t,}a_{t,}unchosen}^{P}\cdot\beta_{Q_{s_{t,}a_{t,}unchosen}^{P}}, Q_{s_{t,}a_{t,}chosen}^{N}\cdot\beta_{Q_{s_{t,}a_{t,}chosen}^{N}}, Q_{s_{t,}a_{t,}unchosen}^{N}\cdot\beta_{Q_{s_{t,}a_{t,}unchosen}^{N}}$, +$\delta_{t}^{P}\cdot\beta_{+\delta_{t}^{P}}$, -$\delta_{t}^{P}\cdot\beta_{-\delta_{t}^{P}}$, +$\delta_{t}^{N}\cdot\beta_{+\delta_{t}^{N}}$, -$\delta_{t}^{N}\cdot\beta_{-\delta_{t}^{N}}$; therefore, parametric regressors described how each learning signal influenced participants’ feelings when presented with an outcome on a given trial. Expected values and prediction errors were derived from individual-level parameter estimates from the hierarchical Bayesian VPRL model estimation^6^. We extracted median Subjective Feeling model coefficient values for each participant from individual-level coefficient posterior distributions. All regressors of interest were z-scored. All motor and visual task measures and six head motion parameters were also included as regressors of no interest.

For each first-level GLM, results for each participant were included into a second-level whole brain analysis of the variance (ANOVA). This allowed us to examine differences in learning and affective signaling between pre-ECT, non-ECT depression, and no-depression groups at visit 1. We performed post-hoc t-tests based on ANOVA results. In addition, first-level GLM results from the VPRL learning signal regressors of interest were included into a second-level one-sample t-test at the whole-brain level where we pooled all participants to determine where separate positive and negative system prediction errors and expected values were tracked in the brain. All statistical analyses were conducted at an uncorrected threshold of p<0.001 in which reported results were selected using a family-wise error (FWE)-corrected threshold of p<0.05 at cluster and peak voxel levels.

**Statistical Analyses**

Hierarchical Bayesian analyses^6^ were performed to estimate posterior distributions of free parameters ($\boldsymbol{\alpha}^{\boldsymbol{P}}$,$\boldsymbol{\alpha}^{\boldsymbol{N}}$**,** $\boldsymbol{\gamma}^{\boldsymbol{P}}$, $\boldsymbol{\gamma}^{\boldsymbol{N}}$, and $\boldsymbol{\tau}$) in the VPRL models of choice behavior and learning (Fig.2A) and subsequently for free parameters in the Subjective Feeling linear regression model ($\boldsymbol{\beta}_{\boldsymbol{constant}}\boldsymbol{,}\boldsymbol{\beta}_{\mathbf{+}\boldsymbol{\delta}_{\boldsymbol{t}}^{\boldsymbol{P}}}\boldsymbol{,}\boldsymbol{\beta}_{\mathbf{-}\boldsymbol{\delta}_{\boldsymbol{t}}^{\boldsymbol{P}}}\boldsymbol{,}\boldsymbol{\beta}_{\mathbf{+}\boldsymbol{\delta}_{\boldsymbol{t}}^{\boldsymbol{N}}}\boldsymbol{,}\boldsymbol{\beta}_{\boldsymbol{-\delta}_{\boldsymbol{t}}^{\boldsymbol{N}}}\boldsymbol{,}\boldsymbol{\beta}_{\boldsymbol{Q}_{\boldsymbol{s}_{\boldsymbol{t,}}\boldsymbol{a}_{\boldsymbol{t,}}\boldsymbol{chosen}}^{\boldsymbol{P}}}\boldsymbol{,}\boldsymbol{\beta}_{\boldsymbol{Q}_{\boldsymbol{s}_{\boldsymbol{t,}}\boldsymbol{a}_{\boldsymbol{t,}}\boldsymbol{unchosen}}^{\boldsymbol{P}}}\boldsymbol{,}\boldsymbol{\beta}_{\boldsymbol{Q}_{\boldsymbol{s}_{\boldsymbol{t,}}\boldsymbol{a}_{\boldsymbol{t,}}\boldsymbol{chosen}}^{\boldsymbol{N}}}\boldsymbol{,}\boldsymbol{\beta}_{\boldsymbol{Q}_{\boldsymbol{s}_{\boldsymbol{t,}}\boldsymbol{a}_{\boldsymbol{t,}}\boldsymbol{unchosen}}^{\boldsymbol{N}}}$; Fig.2B). We discuss evidence of group distributional differences when the 95% highest density interval (HDI) excluded 0 or was within 0.05 of excluding 0^14^. This (arbitrary) threshold acknowledges the limitations of overly strict significance thresholds and allowed us to consider valuable information that may have been missed with stricter cutoffs^14–16^. HDIs are similar to frequentist confidence intervals but describe the most probable 95% of values (as opposed to confidence intervals that describe the probability a value falls within a given range under repeated sampling)^6^. We also visually inspected posterior distributions when assessing group-level differences which provided valuable information, especially regarding the non-normal distribution shapes we observed. We further performed principal components analyses to reduce the dimensionality of both the VPRL parameter set (Fig.2C) and subjective feeling parameter set (Fig.2D).

To investigate individual differences, and to test whether model-based analyses could be used to computationally phenotype individuals without prior information about their clinical diagnosis, we fit VPRL and Subjective Feeling models to each individual in a non-hierarchical Bayesian manner while assuming flat priors. The resulting parameter estimates were concatenated to form a single 14-vector and subjected to PCA (Fig.3A). Finally, we used these putative computational phenotypes in a cross-validated linear discriminant analyses to determine if the 14-vector could be used to classify depression versus no-depression behavior (Fig.3B and Fig.3C). Receiver Operating Characteristic (ROC) analyses with 95% confidence intervals and sensitivity and specificity estimates was performed to determine the accuracy of this approach. For all relevant tests, p<0.05 was deemed significant. Analyses were conducted in R version 4.2.2 and Stan version 2.21.0. (RStan version 2.21.8)^9^.

**eReferences**

1.	Sands LP, Jiang A, Liebenow B, et al. Subsecond fluctuations in extracellular dopamine encode reward and punishment prediction errors in humans. *Sci Adv*. 9(48):eadi4927. doi:10.1126/sciadv.adi4927

2.	Sands LP, Jiang A, Jones RE, Trattner JD, Kishida KT. Valence-partitioned learning signals drive choice behavior and phenomenal subjective experience in humans. Published online March 18, 2023:2023.03.17.533213. doi:10.1101/2023.03.17.533213

3.	Spitzer KK, Williams JBW. Patient Health Questionnaire-9. *APA PsycTests*. doi:https://doi.org/10.1037/t06165-000

4.	Hamilton M. A RATING SCALE FOR DEPRESSION. *J Neurol Neurosurg Psychiatry*. 1960;23(1):56-62. Accessed June 28, 2023. https://www.ncbi.nlm.nih.gov/pmc/articles/PMC495331/

5.	Nasreddine ZS, Phillips NA, Bédirian V, et al. The Montreal Cognitive Assessment, MoCA: A Brief Screening Tool For Mild Cognitive Impairment. *Journal of the American Geriatrics Society*. 2005;53(4):695-699. doi:10.1111/j.1532-5415.2005.53221.x

6.	Kruschke J. *Doing Bayesian Data Analysis: A Tutorial with R, JAGS, and Stan*. Academic Press; 2014.

7.	Kishida KT, Sands LP. A Dynamic Affective Core to Bind the Contents, Context, and Value of Conscious Experience. In: Waugh CE, Kuppens P, eds. *Affect Dynamics*. Springer International Publishing; 2021:293-328. doi:10.1007/978-3-030-82965-0_12

8.	Schurr R, Reznik D, Hillman H, Bhui R, Gershman SJ. Dynamic computational phenotyping of human cognition. *Nat Hum Behav*. 2024;8(5):917-931. doi:10.1038/s41562-024-01814-x

9.	Carpenter B, Gelman A, Hoffman MD, et al. Stan: A Probabilistic Programming Language. *J Stat Softw*. 2017;76:1. doi:10.18637/jss.v076.i01

10.	Johnson AA, Ott MQ, Dogucu M. Chapter 10 Evaluating Regression Models. In: *Bayes Rules! An Introduction to Applied Bayesian Modeling*. Chapman & Hall/CRC texts in statistical science. CRC Press; 2022:255-259. Accessed June 28, 2023. https://www.bayesrulesbook.com/chapter-10.html#cross-validation

11.	Mark Jenkinson 1 , Christian F Beckmann, Timothy E J Behrens, Mark W Woolrich, Stephen M Smith. FSL. *j.neuroimage*. 2012;62(2):782-790. doi:10.1016/j.neuroimage.2011.09.015

12.	Ashburner J, Barnes G, Daunizeau J, et al. SPM12 Manual. https://www.fil.ion.ucl.ac.uk/spm/doc/spm12_manual.pdf

13.	Andersson JLR, Skare S, Ashburner J. How to correct susceptibility distortions in spin-echo echo-planar images: application to diffusion tensor imaging. *Neuroimage*. 2003;20(2):870-888. doi:10.1016/S1053-8119(03)00336-7

14.	Kruschke JK. What to believe: Bayesian methods for data analysis. *Trends in Cognitive Sciences*. 2010;14(7):293-300. doi:10.1016/j.tics.2010.05.001

15.	Ahn WY, Haines N, Zhang L. Revealing Neurocomputational Mechanisms of Reinforcement Learning and Decision-Making With the hBayesDM Package. *Comput Psychiatr*. 2017;1:24-57. doi:10.1162/CPSY_a_00002

16.	Haines N, Vassileva J, Ahn WY. The Outcome-Representation Learning Model: A Novel Reinforcement Learning Model of the Iowa Gambling Task. *Cognitive Science*. 2018;42(8):2534-2561. doi:10.1111/cogs.12688

**eFigure 1: Probabilistic Reward and Punishment with Subjective Rating (PRPwSR) task phases and icon values**

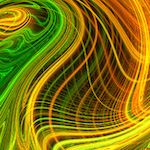

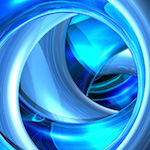

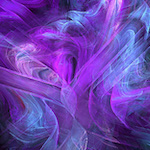

Trial

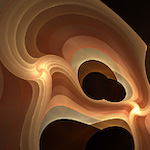

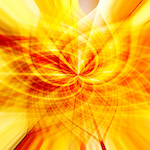

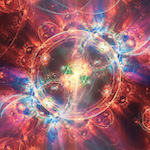

1

25

75

150

0.25(-$1.00) = -$0.25

0.50(-$1.00) = -$0.50

0.75($1.00) = -$0.75

0.25($1.00) = $0.25

0.50($1.00) = $0.50

0.75($1.00) = $0.75

0.75($0.50) = $0.375

0.25($2.50) = $0.625

0.50($1.50) = $0.75

0.25(-$1.25) = -$0.3125

0.50(-$0.75) = -$0.375

0.75(-$1.00) = -$0.1875

Icon value =

outcome probability ($)

**Phase I**

**Phase II**

**Phase III**

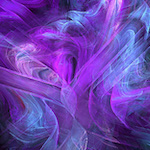

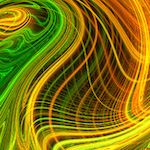

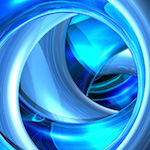

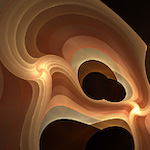

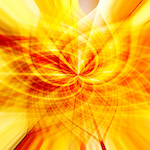

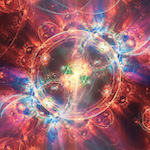

Win Icons

Loss Icons

Example of six fractal images that are presented throughout the PRPwSR task. Icon expected values are the product of the icon probability and dollar amount presented (e.g., purple icon in phase 1: 0.75 probability of $1 = $0.75 value). Phase I includes 3 win icons with fixed probabilities of earning $1 (versus $0). Phase II introduces three loss icons with fixed probabilities of losing $1 (versus $0). In phase III, icon probabilities remain the same, but associated dollar amounts change. This changes icon expected values (e.g., purple icon in phase 1 and 2 displays $1 but in phase 3 displays $0.50; expected value changes from $0.75 to $0.375).

**eFigure 2: Expected versus actual icon value for each cohort**

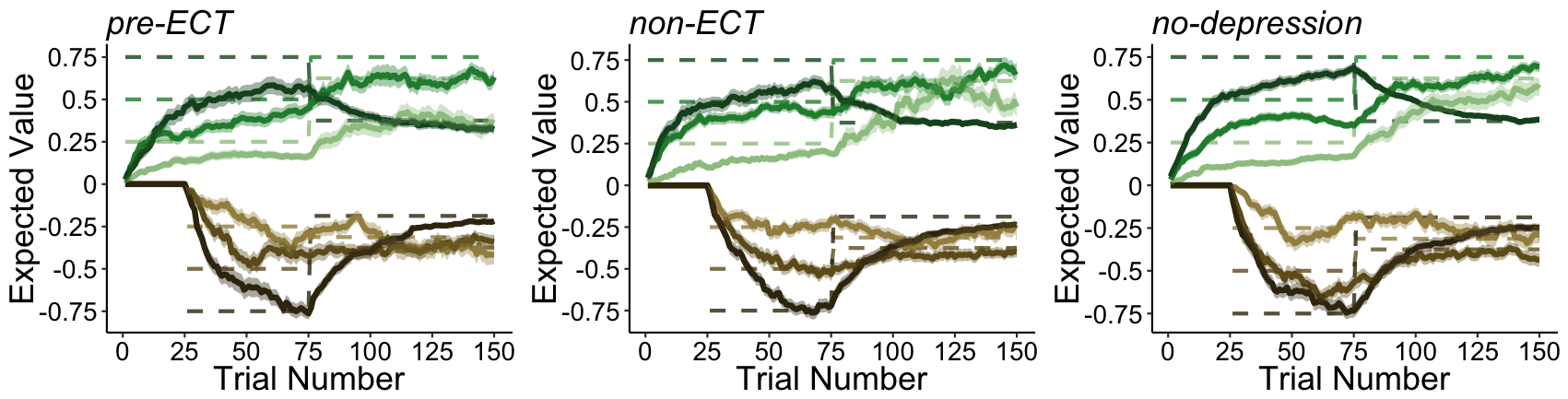

**A**

**B**

**C**

Learned versus actual expected values for each of the six icon groups throughout all 150 trials of the PRPwSR task for **A)** pre-ECT, **B)** non-ECT, and **C)** no-depression groups. Dashed lines indicate true icon expected values, which switch at phase III (trial 75). Solid lines with $\pm$ 1 SEM indicate participants’ learned expected icon values throughout the task. Win icons are represented in green and loss icons are represented in brown.

**eFigure 3: PRPwSR task performance metrics by cohort**

Monetary Gains

Monetary Losses

**A**

Total Monetary Outcome

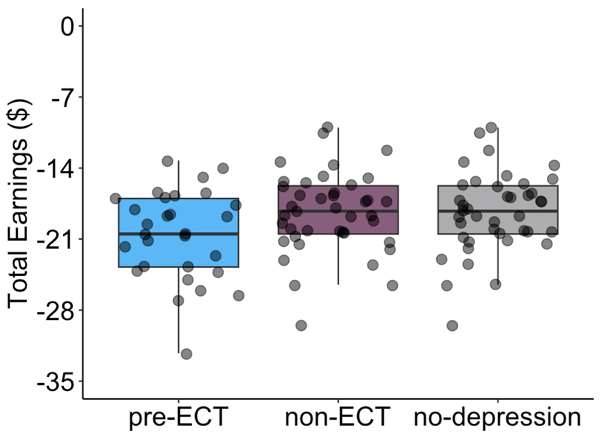

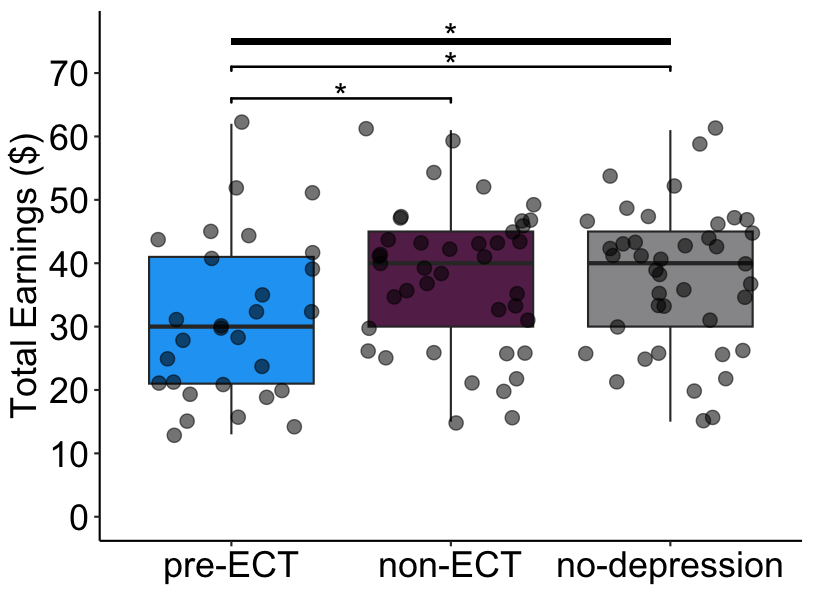

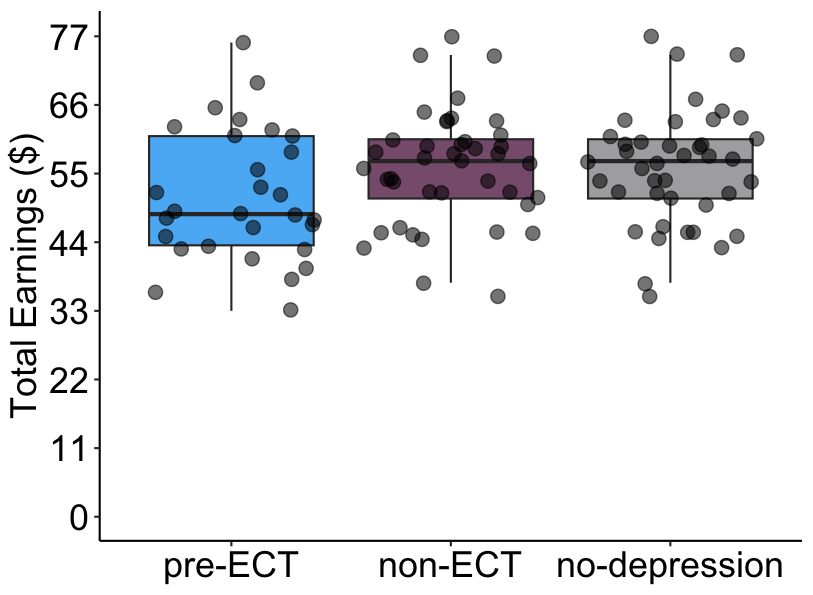

**B**

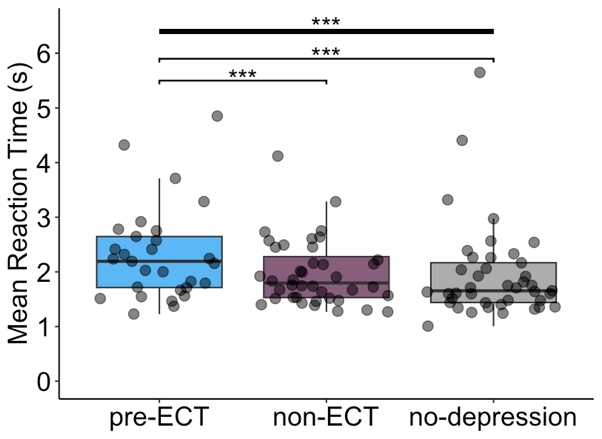

Cohort distributions of total earnings and mean reaction times during the PRPwSR task. One-way ANOVA with tukey’s post-hoc tests were performed for each comparison plotted. **A)** left – total monetary outcome from the PRPwSR task (F=3.75, p=0.03; pre-ECT vs. non-ECT p-adj=0.04, pre-ECT vs. no-depression p-adj=0.04); middle – monetary gains from win icons only; right – monetary losses from loss icons only. **B)** Mean reaction time measured as the time in seconds between icon presentation and option selection (F=47.54, p=2.60e-21; pre-ECT vs. non-ECT p-adj=2.68e-8, pre-ECT vs. no-depression p-adj=2.68e-8).

**eFigure 4. Model-predicted versus participant-reported ratings**

**A**

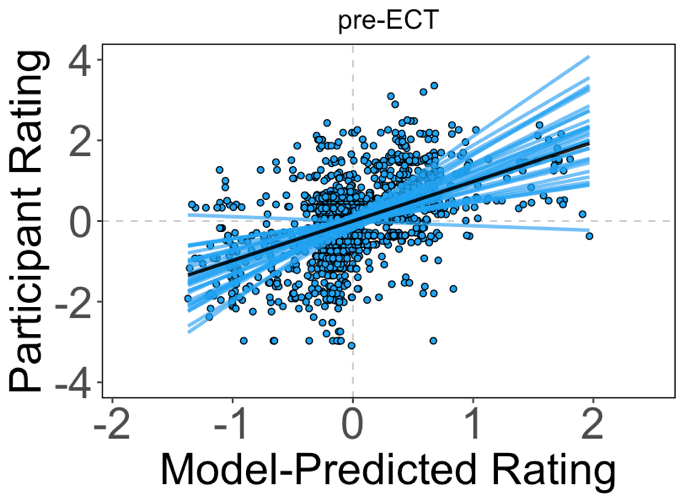

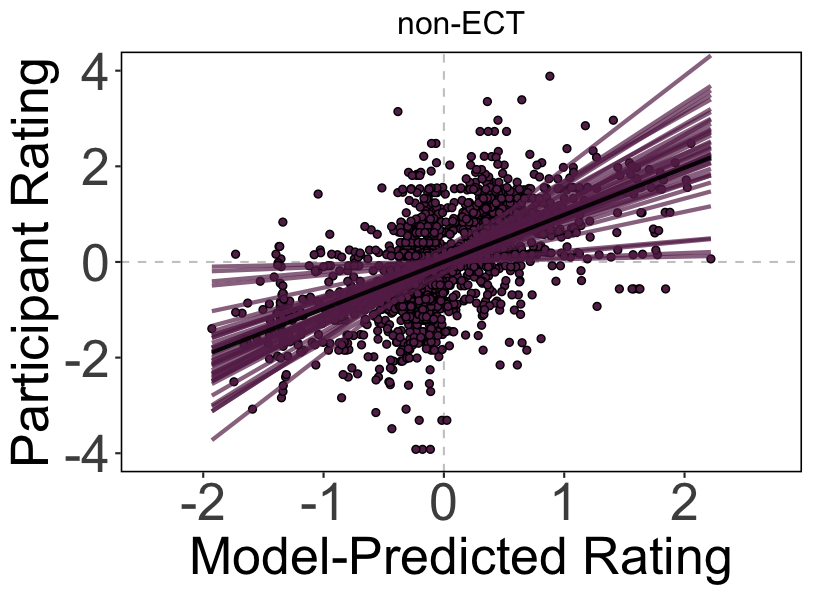

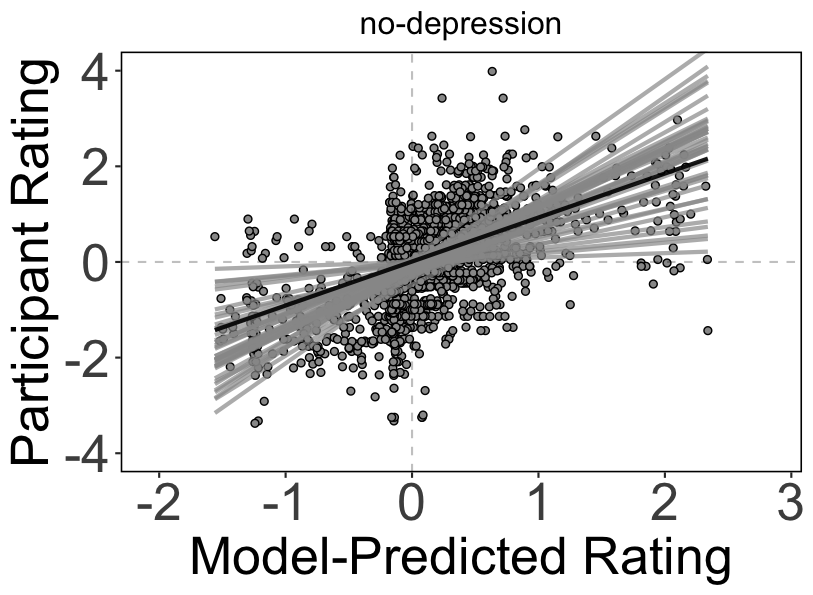

**B**

**C**

In the PRPwSR task, participants completed 50 rating trials regarding how they felt about their last monetary outcome. We derived expected values and prediction errors from these rating trails for each participant and used these as independent variables in a Bayesian linear regression model to predict participant ratings. Model-predicted versus actual participant ratings are displayed for **(A)** pre-ECT (R^2^=0.272, p=0.0374), **(B)** non-ECT (R^2^=0.357, p=0.0619), and **(C)** no-depression (R^2^=0.348, p=0.0496).

**eFigure 5. Pairs plots of individually-fit learning parameter and subjective rating coefficient PCA**

no-depression

pre-ECT

non-ECT

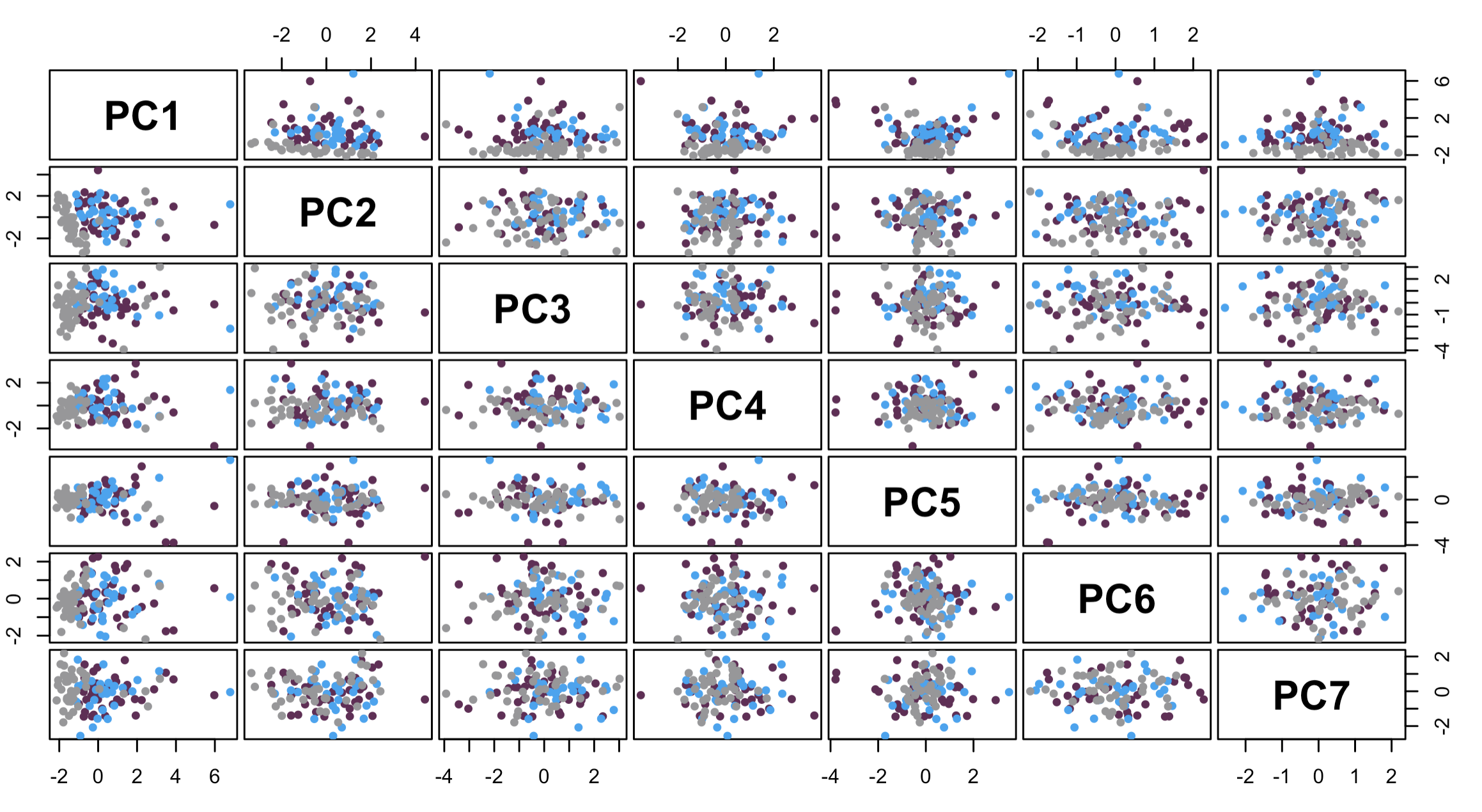

no-depression

pre-ECT

non-ECT

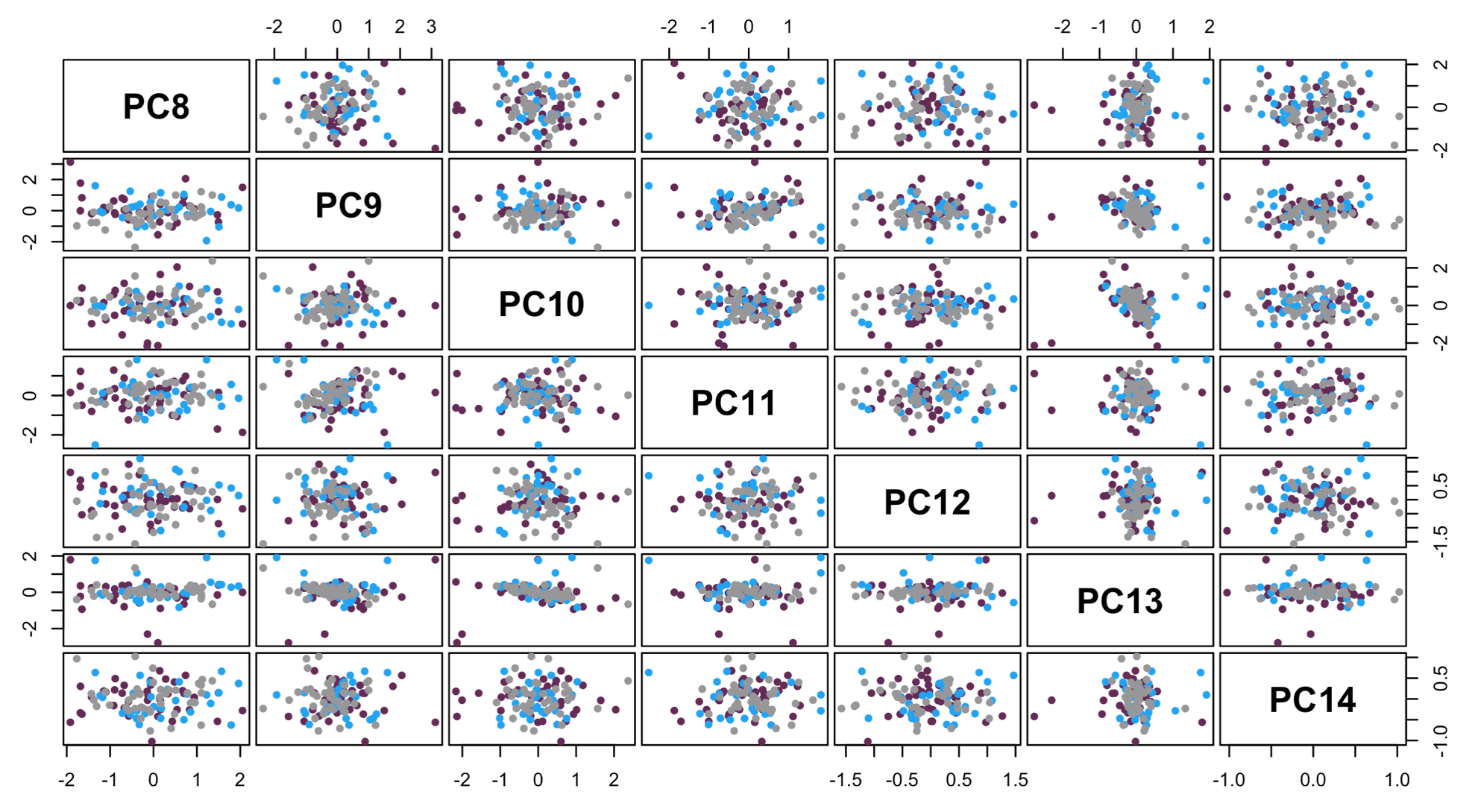

Pairs plots of principal components 1-14 from principal component analysis using participants’ VPRL learning parameters and Subjective Feeling model coefficients. For each graph, pre-ECT (blue), non-ECT (purple), and no-depression (gray) groups are depicted. See Fig.3A in main text for information about principal component 1 & 2 coefficients.

| **eTable 1. Participant Demographic and Clinical Characteristics** | | | | | |
| --- | --- | --- | --- | --- | --- |
| **Variable** | **pre-ECT**  **(n = 29)** | **non-ECT**  **(n = 40)** | **no-depression**  **(n = 41)** | **Group Comparison** | **Post-hoc comparisons** |
| Age, mean (SD), years | 39.07 (14.92) | 42.43 (15.11) | 40.10 (15.20) | F_2,92_ = 0.46 (ns) |  |
| Gender: |  |  |  | $\chi_{4}^{2}=$ 4.02 (ns) |  |
| Male | 12 | 12 | 14 |  |  |
| Female | 16 | 28 | 27 |  |  |
| Non-binary | 1 | 0 | 0 |  |  |
| Race: |  |  |  | $\chi_{8}^{2}=$ 10.30 (ns) |  |
| AIAN | 1 | 0 | 0 |  |  |
| Asian | 0 | 1 | 5 |  |  |
| Black/African American | 3 | 4 | 5 |  |  |
| White/Caucasian | 25 | 35 | 31 |  |  |
| Other | 1 | 0 | 1 |  |  |
| Ethnicity: |  |  |  | $\chi_{2}^{2}=$ 2.64 (ns) |  |
| Hispanic/Latino | 2 | 2 | 0 |  |  |
| Not Hispanic/Latino | 27 | 38 | 41 |  |  |
| Clinical Scores: |  |  |  |  |  |
| MOCA | 26.83 $\pm$2.56 | 26.38$\pm$ 3.42 | 27.0 $\pm$2.02 | F_2,107_ = 0.578 (ns) |  |
| PHQ-9 | 19.86 $\pm$ 5.49 | 11.40 $\pm$6.52 | 1.66 $\pm$ 2.49 | F_2,107_ = 112^a^ | pre-ECT > non-ECT^a^ |
|  |  |  |  |  | pre-ECT > no-depression^a^ |
|  |  |  |  |  | non-ECT > no-depression^a^ |
| HAM-D | 24.69 $\pm$ 6.48 | 13.45$\pm$8.09 | 1.98 $\pm$2.13 | F_2,107_ = 121.5^a^ | pre-ECT > non-ECT^a^ |
|  |  |  |  |  | pre-ECT > no-depression^a^ |
|  |  |  |  |  | non-ECT > no-depression^a^ |
| Abbreviations: ns: not significant; AIAN: American Indian and Alaska Native; MOCA: Montreal Cognitive Assessment; PHQ-9: Patient Health Questionnaire-9; HAM-D: Hamilton Depression Rating Scale. ^a^p<0.001 (post-hoc Tukey HSD). | | | | | |

| **eTable 2. Current psychiatric medications and comorbidities reported by participants with depression** | | |
| --- | --- | --- |
|  | pre-ECT | non-ECT |
| Sample Size | 29 | 40 |
| Medications: |  |  |
| Antidepressant (SSRI; SNRI; NDRI; SARI) | 23 | 30 |
| Antipsychotic/Anticonvulsant/Mood Stabilizer | 18 | 10 |
| Anxiolytic/Hypnotic (benzodiazepine; antihistamine) | 11 | 10 |
| Stimulant | 4 | 5 |
| None Reported | 0 | 8 |
| Comorbidities: |  |  |
| Anxiety/Panic Disorder | 21 | 17 |
| Attention-Deficit/Hyperactivity Disorder | 7 | 8 |
| Bipolar Disorder | 7 | 6 |
| Borderline or Dissociative Personality Disorder | 6 | 2 |
| Eating Disorder | 2 | 2 |
| Obsessive Compulsive Disorder | 1 | 3 |
| Post-Traumatic Stress Disorder | 7 | 1 |
| Psychosis/Schizophrenia/Schizoaffective Disorder | 1 | 1 |
| Sleep Disorder | 3 | 0 |
| None Reported | 5 | 13 |
| Note: Many subjects report multiple medication types and multiple comorbidities (data reported as count). | | |

| **eTable 3. Performance metrics of different group-level models** | | | |
| --- | --- | --- | --- |
|  |  | *Model evidence* | *Predictive density* |
| Model 1 |  |  |  |
| all participants |  | -9076.45 | -9486.40 |
| Model 2 |  |  |  |
| pre-ECT & non-ECT |  | -5830.33 | -6081.5 |
| no-depression |  | -3244.60 | -3404.9 |
| sum |  | -9074.93 | -9486.40 |
| Model 3 |  |  |  |
| pre-ECT |  | -2481.18 | -2586.40 |
| non-ECT |  | -3345.45 | -3492.40 |
| no-depression |  | -3244.60 | -3404.9 |
| sum |  | **-9071.22** | **-9483.70** |
| We fit the VPRL model using hierarchical Bayesian inference to three models under different group-level assumptions. We compared all models via estimates of the marginal likelihood and posterior predictive density of the VPRL model by summing these criteria across models for each group-level model assumption. The models with the maximum (least negative) criteria values are preferred (bolded). | | | |

| **eTable 4. Comparison of VPRL model parameter posterior distributions across cohorts** | | | | | | | | | | | | | | | | | | | |
| --- | --- | --- | --- | --- | --- | --- | --- | --- | --- | --- | --- | --- | --- | --- | --- | --- | --- | --- | --- |
|  |  |  |  |  |  |  | |  | *Group Comparisons* | | | | | | | | | | |
| Learning Parameter |  | *pre-ECT*  *(n = 29)* |  | *non-ECT*  *(n = 40)* |  | | *no-depression*  *(n = 41)* | | |  | *non-ECT –*  *pre-ECT* | |  | *no-depression –*  *pre-ECT* | |  | | *no-depression – non-ECT* | |
|  |  | Median  [95% HDI] |  | Median  [95% HDI] |  | | Median  [95% HDI] | | |  | Median Difference  [95% HDI] | % Credible differences |  | Median Difference  [95% HDI] | % Credible differences |  | Median Difference  [95% HDI] | | % Credible differences |
| $\alpha^{P}$ |  | **0.14**  **[0.08,**  **0.21]** |  | **0.20**  **[0.14,**  **0.26]** |  | | **0.16**  **[0.13,**  **0.19]** | | |  | **0.06**  **[-0.04, 0.15]** | [12.51, 87.49] |  | 0.02  [-0.06, 0.09] | [33.86, 66.14] |  | **-0.04**  **[-0.11, 0.03]** | | [88.41, 11.59] |
| $\gamma^{P}$ |  | **0.76**  **[0.52,**  **1]** |  | **0.83**  **[0.52,**  **1]** |  | | **0.83**  **[0.57,**  **1]** | | |  | 0.05  [-0.37, 0.42] | [40.90, 59.10] |  | 0.05  [-0.34, 0.41] | [40.01, 59.99] |  | 0.003  [-0.38, 0.41] | | [49.46, 50.54] |
| $\alpha^{N}$ |  | **0.36**  **[1.17e^-5^, 0.79]** |  | **0.46**  **[1.76e^-4^,**  **0.66]** |  | | **0.54**  **[0.03,**  **0.77]** | | |  | 0.04  [-0.57, 0.58] | [45.27, 54.73] |  | 0.13  [-0.50, 0.70] | [35.39, 64.61] |  | 0.08  [-0.48, 0.62] | | [37.16, 62.84] |
| $\gamma^{N}$ |  | **0.11**  **[4.07e^-4^, 0.49]** |  | **0.09**  **[2.62e^-5^, 0.53]** |  | | **0.18**  **[0.03,**  **0.47]** | | |  | -0.02  [-0.50, 0.49] | [56.55, 43.45] |  | 0.06  [-0.38, 0.43] | [33.80, 66.20] |  | 0.08  [-0.42, 0.46] | | [25.34, 74.66] |
| $1/\tau$ |  | **0.09**  **[0.05,**  **0.18]** |  | **0.16**  **[0.05,**  **0.25]** |  | | **0.11**  **[0.05,**  **0.19]** | | |  | 0.05  [-0.08, 0.19] | [23.91, 76.09] |  | 0.02  [-0.10, 0.13] | [38.56, 61.44] |  | -0.04  [-0.17, 0.11] | | [69.24, 30.76] |
| Abbreviations: HDI: highest density interval. Bold font indicates evidence of distributional significance. | | | | | | | | | | | | | | | | | | | |

| **eTable 5. Positive and negative system prediction error and expected value results from whole-brain one-sample t-test pooling all participants** | | | | | | |
| --- | --- | --- | --- | --- | --- | --- |
| Parameter | Region | Voxels | Cluster-level p-value | Peak-level p-value | Statistic | Peak MNI coordinates  [x y z] |
| **+**$\boldsymbol{\delta}_{\boldsymbol{t}}^{\boldsymbol{P}}$ | Right caudate | 162 | **1.14e-12** | **2.48e-9** | T(103) = 8.76 | [6 10 -2] |
|  | Left putamen | 39 | **3.87e-6** | **1.09e-6** | T(103) = 7.55 | [-14 14 0] |
|  | Right inferior frontal operculum | 101 | **8.63e-10** | **4.86e-6** | T(103) = 7.24 | [46 10 24] |
|  | Left inferior frontal gyrus | 10 | **0.001** | **1.15e-4** | T(103) = 6.59 | [-44 42 10] |
|  | Right middle frontal gyrus | 56 | **2.91e-7** | **0.001** | T(103) = 6.23 | [40 28 22] |
|  | Right middle cingulate cortex | 30 | **1.78e-5** | **0.001** | T(103) = 6.21 | [4 2 30] |
|  | Left anterior cingulate cortex | 31 | **1.49e-5** | **0.001** | T(103) = 6.14 | [-2 44 -4] |
|  | Right insula | 21 | **9.59e-5** | **0.001** | T(103) = 6.06 | [32 22 -2] |
|  | Left middle cingulate cortex | 33 | **1.05e-5** | **0.003** | T(103) = 5.87 | [-4 -32 38] |
|  | Left insula | 22 | **7.86e-5** | **0.003** | T(103) = 5.86 | [-30 20 -4] |
|  | Left inferior temporal gyrus | 42 | **2.39e-6** | **0.005** | T(103) = 5.70 | [-44 -58 -10] |
|  | Right supramarginal gyrus | 15 | **3.39e-4** | **0.007** | T(103) = 5.63 | [50 -30 44] |
|  | Left anterior cingulate cortex | 11 | **0.001** | **0.016** | T(103) = 5.42 | [-4 36 12] |
|  | Left inferior parietal lobule | 23 | **6.47e-5** | **0.017** | T(103) = 5.40 | [-32 -56 46] |
| **-**$\boldsymbol{\delta}_{\boldsymbol{t}}^{\boldsymbol{P}}$ |  |  |  |  | NS |  |
| $\boldsymbol{Q}_{\boldsymbol{s}_{\boldsymbol{t,}}\boldsymbol{a}_{\boldsymbol{t,}}\boldsymbol{chosen}}^{\boldsymbol{P}}$ | Left medial prefrontal cortex | 17 | **2.58e-4** | **0.006** | T(103) = 5.66 | [-6 56 18] |
|  | Left posterior cingulate cortex | 25 | **5.48e-5** | **0.011** | T(103) = 5.51 | [-8 -48 30] |
|  | Right middle occipital gyrus | 184 | **2.93e-13** | **9.43e-7** | T(103) = -7.58 | [34 -86 10] |
|  | Left middle occipital gyrus | 140 | **2.18e-11** | **4.03e-6** | T(103) = -7.28 | [-24 -94 10] |
|  | Left fusiform gyrus | 164 | **1.98e-12** | **4.87e-5** | T(103) = -6.77 | [-40 -64 -14] |
|  | Left supplementary motor area | 123 | **1.30e-10** | **8.80e-5** | T(103) = -6.64 | [-4 20 48] |
|  | Right superior parietal lobule | 207 | **3.53e-14** | **1.76e-4** | T(103) = -6.50 | [32 -62 52] |
| Parameter | Region | Voxels | Cluster-level p-value | Peak-level p-value | Statistic | Peak MNI coordinates  [x y z] |
|  | Right fusiform gyrus | 73 | **4.36e-8** | **1.85e-4** | T(103) = -6.48 | [28 -54 -14] |
|  | Left inferior parietal lobule | 35 | **9.80e-6** | **0.001** | T(103) = -6.08 | [-30 -56 54] |
|  | Right insula | 26 | **4.57e-5** | **0.003** | T(103) = -5.87 | [34 20 -2] |
|  | Left inferior frontal gyrus | 12 | **0.001** | **0.009** | T(103) = -5.55 | [-42 24 30] |
|  | Left precentral gyrus | 13 | **0.001** | **0.010** | T(103) = -5.52 | [-42 6 36] |
| $\boldsymbol{Q}_{\boldsymbol{s}_{\boldsymbol{t,}}\boldsymbol{a}_{\boldsymbol{t,}}\boldsymbol{unchosen}}^{\boldsymbol{P}}$ |  |  |  |  | NS |  |
| **+**$\boldsymbol{\delta}_{\boldsymbol{t}}^{\boldsymbol{N}}$ | Right supplementary motor area | 70 | **1.11e-7** | **3.71e-6** | T(103) = 7.30 | [8 14 64] |
|  | Left insula | 253 | **2.33e-15** | **1.88e-5** | T(103) = 6.97 | [-38 22 -4] |
|  | Right insula | 94 | **6.55e-9** | **4.04e-4** | T(103) = 6.31 | [40 20 -8] |
|  | Left inferior temporal gyrus | 17 | **3.20e-4** | **4.61e-4** | T(103) = 6.28 | [-42 -54 -8] |
|  | Right inferior frontal operculum | 81 | **2.93e-8** | **5.72e-7** | T(103) = 5.86 | [44 12 36] |
|  | Right middle cingulate cortex | 14 | **0.001** | **0.012** | T(103) = 5.71 | [8 24 38] |
|  | Left precentral gyrus | 10 | **0.001** | **0.028** | T(103) = 5.33 | [-42 6 34] |
| **-**$\boldsymbol{\delta}_{\boldsymbol{t}}^{\boldsymbol{N}}$ |  |  |  |  | NS |  |
| $\boldsymbol{Q}_{\boldsymbol{s}_{\boldsymbol{t,}}\boldsymbol{a}_{\boldsymbol{t,}}\boldsymbol{chosen}}^{\boldsymbol{N}}$ |  |  |  |  | NS |  |
| $\boldsymbol{Q}_{\boldsymbol{s}_{\boldsymbol{t,}}\boldsymbol{a}_{\boldsymbol{t,}}\boldsymbol{unchosen}}^{\boldsymbol{N}}$ |  |  |  |  | NS |  |
| Analyses were performed using FWE-corrected threshold of p<0.05. Bold indicates significance. NS – not significant. | | | | | | |

| **eTable 6. Comparison of Subjective Feeling model coefficient posterior distributions across cohorts** | | | | | | | | | | | | | | | | | | |
| --- | --- | --- | --- | --- | --- | --- | --- | --- | --- | --- | --- | --- | --- | --- | --- | --- | --- | --- |
|  |  |  |  |  |  |  |  | *Group Comparisons* | | | | | | | | | | |
| Coefficient |  | *pre-ECT*  *(n = 29)* |  | *non-ECT*  *(n = 40)* |  | *no-depression*  *(n = 41)* | | |  | *non-ECT –*  *pre-ECT* | |  | *no-depression –*  *pre-ECT* | | |  | *no-depression –*  *non-ECT* | |
|  |  | Median  [95% HDI] |  | Median  [95% HDI] |  | Median  [95% HDI] | | |  | Median Difference  [95% HDI] | % Credible differences | |  | Median Difference  [95% HDI] | % Credible differences |  | Median Difference  [95% HDI] | % Credible differences |
| $\beta_{+\delta_{t}^{P}}$ |  | **0.80**  **[0.67,**  **0.93]** |  | **0.74**  **[0.65,**  **0.84]** |  | **0.89**  **[0.76,**  **1.01]** | | |  | -0.05  [-0.21,  0.11] | [74.87, 25.13] | |  | 0.09  [-0.09,  0.27] | [17.0,  83.0] |  | **0.14**  **[-0.01,**  **0.30]** | [3.76,  96.24] |
| $\beta_{-\delta_{t}^{P}}$ |  | **-0.35**  **[-0.58,**  **-0.12]** |  | **-0.47**  **[-0.63,**  **-0.31]** |  | -0.06  [-0.30,  0.17] | | |  | -0.12  [-0.40,  0.16] | [79.43, 20.57] | |  | **0.29**  **[-0.05,**  **0.61]** | [4.57,  95.43] |  | **0.41**  **[0.12,**  **0.69]** | [0.27,  99.73] |
| $\beta_{Q_{s_{t,}a_{t,}chosen}^{P}}$ |  | **1.14**  **[0.74,**  **1.55]** |  | **1.06**  **[0.80,**  **1.33]** |  | **0.90**  **[0.53,**  **1.26]** | | |  | -0.08  [-0.56,  0.39] | [63.22, 36.78] | |  | -0.25  [-0.79,  0.29] | [81.64, 18.36] |  | -0.17  [-0.61,  0.28] | [76.69, 23.31] |
| $\beta_{Q_{s_{t,}a_{t,}unchosen}^{P}}$ |  | -0.65  [-1.35,  0.06] |  | **-0.66**  **[-1.04,**  **-0.27]** |  | 0.30  [-0.36,  0.96] | | |  | -0.004  [-0.81,  0.80] | [50.44, 49.56] | |  | **0.95**  **[-0.01,**  **1.91]** | [2.64,  97.36] |  | **0.96**  **[0.20,**  **1.72]** | [0.67,  99.33] |
| $\beta_{+\delta_{t}^{N}}$ |  | **-0.89**  **[-1.11,**  **-0.68]** |  | **-1.23**  **[-1.40,**  **-1.05]** |  | -**1.11**  **[-1.36,**  **-0.87]** | | |  | -**0.34**  **[-0.61,**  **-0.06]** | [0.004,  0.96] | |  | -0.22  [-0.55,  0.11] | [90.65,  9.35] |  | 0.11  [-0.18,  0.41] | [22.55, 77.45] |
| $\beta_{-\delta_{t}^{N}}$ |  | **0.25**  **[-0.04,**  **0.55]** |  | 0.15  [-0.10,  0.40] |  | 0.25  [-0.06,  0.56] | | |  | -0.10  [-0.49,  0.29] | [69.36, 30.64] | |  | -0.003  [-0.43,  0.43] | [50.63, 49.37] |  | 0.10  [-0.30,  0.50] | [31.66, 68.34] |
| $\beta_{Q_{s_{t,}a_{t,}chosen}^{N}}$ |  | -1.26  [-2.76,  0.27] |  | **-1.88**  **[-3.22,**  **-0.54]** |  | -1.47  [-3.11,  0.11] | | |  | -0.64  [-2.64,  1.40] | [73.23, 26.77] | |  | -0.23  [-2.45,  1.96] | [58.22, 41.78] |  | 0.41  [-1.68,  2.50] | [35.03, 64.97] |

|  |  |  |  |  |  |  |  | *Group Comparisons* | | | | | | | | | | |
| --- | --- | --- | --- | --- | --- | --- | --- | --- | --- | --- | --- | --- | --- | --- | --- | --- | --- | --- |
| Coefficient |  | *pre-ECT*  *(n = 29)* |  | *non-ECT*  *(n = 40)* |  | *no-depression*  *(n = 41)* | | |  | *non-ECT –*  *pre-ECT* | |  | *no-depression –*  *pre-ECT* | | |  | *no-depression –*  *non-ECT* | |
|  |  | Median  [95% HDI] |  | Median  [95% HDI] |  | Median  [95% HDI] | | |  | Median Difference  [95% HDI] | % Credible differences | |  | Median Difference  [95% HDI] | % Credible differences |  | Median Difference  [95% HDI] | % Credible differences |
| $\beta_{Q_{s_{t,}a_{t,}unchosen}^{N}}$ |  | -0.30  [-1.33,  0.75] |  | 0.32  [-0.47,  1.11] |  | **1.75**  **[0.55,**  **2.87]** | | |  | 0.63  [-0.67,  1.96] | [17.43, 82.57] | |  | **2.05**  **[0.48,**  **3.59]** | [0.62,  99.38] |  | **1.43**  **[0.03,**  **2.84]** | [2.56,  97.44] |
| Abbreviations: HDI: highest density interval. Bold font indicates evidence of distributional significance. | | | | | | | | | | | | | | | | | | |

| **eTable 7. Predicted versus actual depression status from leave-one-out cross-validation linear discriminant analysis** | | | |
| --- | --- | --- | --- |
|  | *Predicted* | |  |
| *Actual* | Depression | No Depression | Total |
| Depression | 59 | 6 | 65 |
| No Depression | 10 | 34 | 44 |
| Total | 69 | 40 | 109 |
